# Identifying risk factors for COVID-19 severity and mortality in the UK Biobank

**DOI:** 10.1101/2021.05.10.21256935

**Authors:** Iqbal Madakkatel, Catherine King, Ang Zhou, Anwar Mulugeta, Amanda Lumsden, Mark McDonnell, Elina Hyppönen

## Abstract

Severe acute respiratory syndrome coronavirus has infected over 114 million people worldwide as of March 2021, with worldwide mortality rates ranging between 1-10%. We use information on up to 421,111 UK Biobank participants to identify possible predictors for long-term susceptibility to severe COVID-19 infection (*N* =1,088) and mortality (*N* =376). We include 36,168 predictors in our analyses and use a gradient boosting decision tree (GBDT) algorithm and feature attribution based on Shapley values, together with traditional epidemiological approaches to identify possible risk factors. Our analyses show associations between socio-demographic factors (e.g. age, sex, ethnicity, education, material deprivation, accommodation type) and lifestyle indicators (e.g. smoking, physical activity, walking pace, tea intake, and dietary changes) with risk of developing severe COVID-19 symptoms. Blood (cystatin C, C-reactive protein, gamma glutamyl transferase and alkaline phosphatase) and urine (microalbuminuria) biomarkers measured more than 10 years earlier predicted severe COVID-19. We also confirm increased risks for several pre-existing disease outcomes (e.g. lung diseases, type 2 diabetes, hypertension, circulatory diseases, anemia, and mental disorders). Analyses on mortality were possible within a sub-group testing positive for COVID-19 infection (*N* =1,953) with our analyses confirming association between age, smoking status, and prior primary diagnosis of urinary tract infection.

**SUMMARY:** Our hypothesis-free approach combining machine learning with traditional epidemiological methods finds a number of risk factors (sociodemographic, lifestyle, and psychosocial factors, biomarkers, disease outcomes and treatments) associated with developing severe COVID-19 symptoms and COVID-19 mortality.

## Background

Severe acute respiratory syndrome coronavirus (SARS-CoV-2) has infected over 114 million people worldwide as of March 2021, with worldwide mortality rates ranging between 1-10%. While many patients will have a mild or even asymptomatic infection, 10-20% of patients experience severe infection requiring hospitalization. In critically ill patients the so called ‘cytokine storm’ characterized by excessive production of proinflammatory molecules can lead to multi-organ damage particularly in the lungs manifesting as acute respiratory distress syndrome, as well as disseminated intravascular coagulation and shock, and is associated with high levels of mortality [1]. In addition, survivors of severe coronavirus disease (COVID-19) are likely to have long-term adverse health effects [2, 3]. Characterizing those at risk of severe infection and mortality, can inform public health strategies to prevent and manage the pandemic, and provide insights into the risk factors reflecting longer-term susceptibility to severe infection.

Machine learning (ML) is the application of computer algorithms which learn from data. While traditional statistical testing requires assumptions and *a priori* knowledge, ML has the advantage of being hypothesis free (i.e., not requiring *a priori* assumptions on causality). It is also able to handle large complex datasets. Previous studies have used ML to explore various aspects relating to the diagnosis and prognosis of COVID-19 disease, including approaches to treatment and management, forecasting, and anti-viral drug discovery [4-6]. In this study, we use ML to explore characteristics reflecting longer-term susceptibility to infection. We use information from over 30,000 features which have been collected up to 14 years before the COVID-19 crisis, with an aim to identify characteristics associated with the severity and/or mortality from COVID-19. We use a novel approach where ‘risk factor discovery’ is conducted using ML, followed by standard epidemiological analyses to facilitate confounder adjustments and interpretation [7]. Our study is based on information from the UK Biobank [8, 9]. Established in 2007, it includes >500,000 participants and is one of the world’s largest and most comprehensive prospective cohort studies, enabling us to examine the possible contribution of an extensive range of potential risk factors and biomarkers. We use an ML algorithm called gradient boosting decision trees (GBDT) [10] and conduct further epidemiological analyses to explore the importance and quantify the effects of the identified COVID-19 predictors.

## Methods

### Participants

The UK Biobank contains genetic, physical, and clinical data on over 500,000 middle to older aged participants (aged 37-73 years) recruited between March 13, 2006 and October 1, 2010 from England (89%), Scotland (7%) and Wales (4%) through 22 assessment centers and followed up by linkage to hospital, cancer and mortality registrations and online surveys. COVID-19 test result data up to July 26, 2020 for the participants from England were provided by Public Health England [11] and accordingly, our study was confined to the participants from England. Participants who died before January 2020 were also excluded from our analyses. COVID-19 diagnosis was made based on a positive reverse transcription-polymerase chain reaction (RT-PCR) test. Severe COVID-19 infection was defined by hospital admission with diagnosis or death under ICD 10 codes U07.1 and U07.2 (recorded up to May 31 and June 28, 2020, respectively). For severe COVID-19 analyses, the control group consisted of participants living in England, excluding those who had received a positive COVID-19 test. In mortality analyses, the control group consisted of participants who tested positive and/or had COVID-19 disease requiring hospitalization.

In this study, we considered all information collected at the baseline assessment using touchscreen questionnaires, biomarker profiling, and results from clinical examinations, in addition to disease coding derived from linkage to cancer registrations (up to December 2016) and hospital episode statistics (HES) (up to September 2019). As the data was not sufficiently structured for our analyses, we ran an automated pre-processing step using a specifically designed software package for UK Biobank, PHESANT (PHEnome Scan Analysis [12] (Supplementary Methods), available in R. We removed baseline features which were recorded for less than 90% of the participants. Information obtained from online follow-up surveys or sub-samples of the cohort were excluded from our analyses due to their low coverage. Supplementary Table 1 and Supplementary Table 2 show UK Biobank variables included in our severity and mortality analyses, respectively. The pre-processing resulted in 36,168 features for severe COVID-19 and 36,145 features for COVID-19 mortality analyses, with 92% of the features representing HES and cancer linkage data. Supplementary Table 3 shows category-wise counts of those features for severity and mortality analyses.

**Table 1 :**
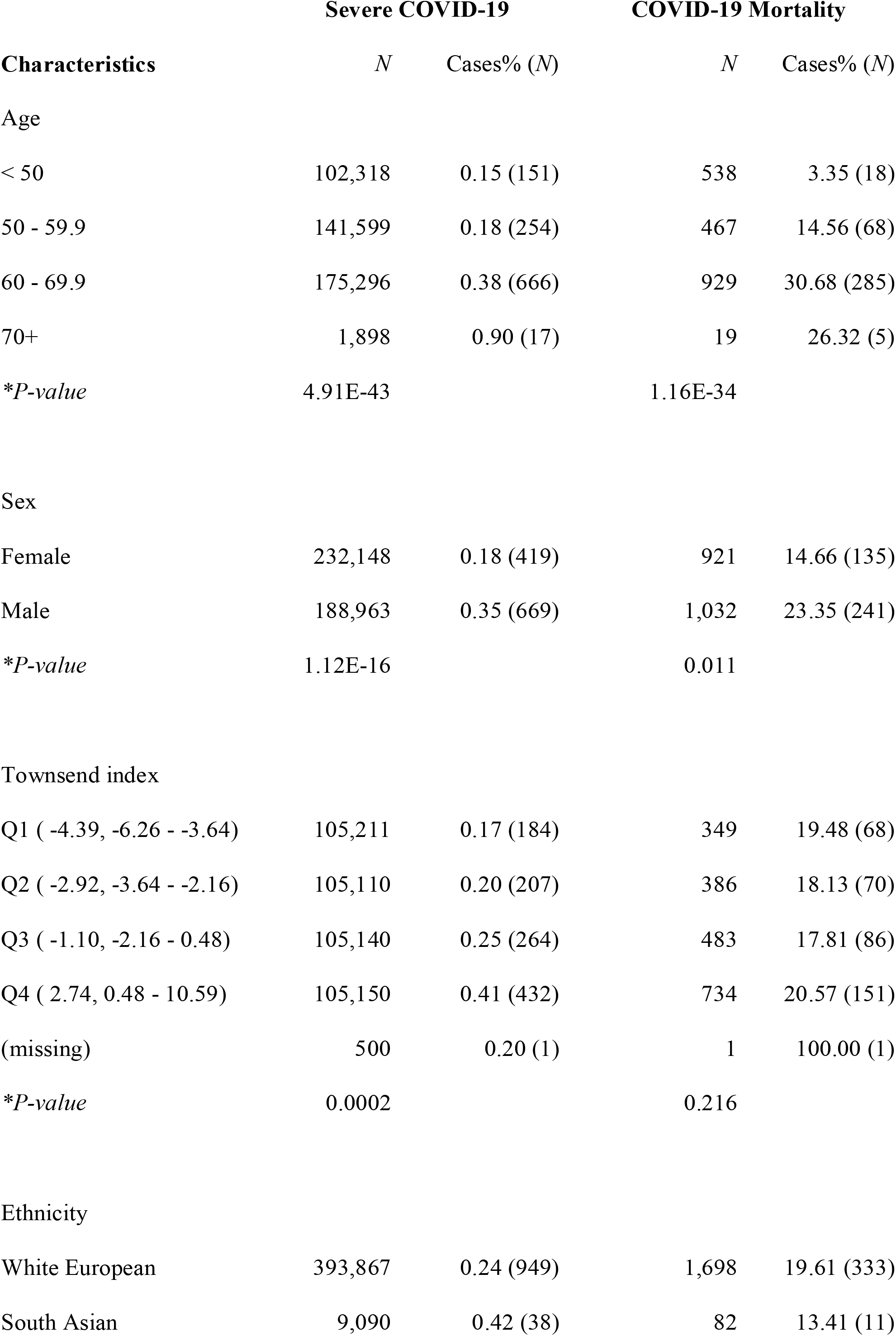
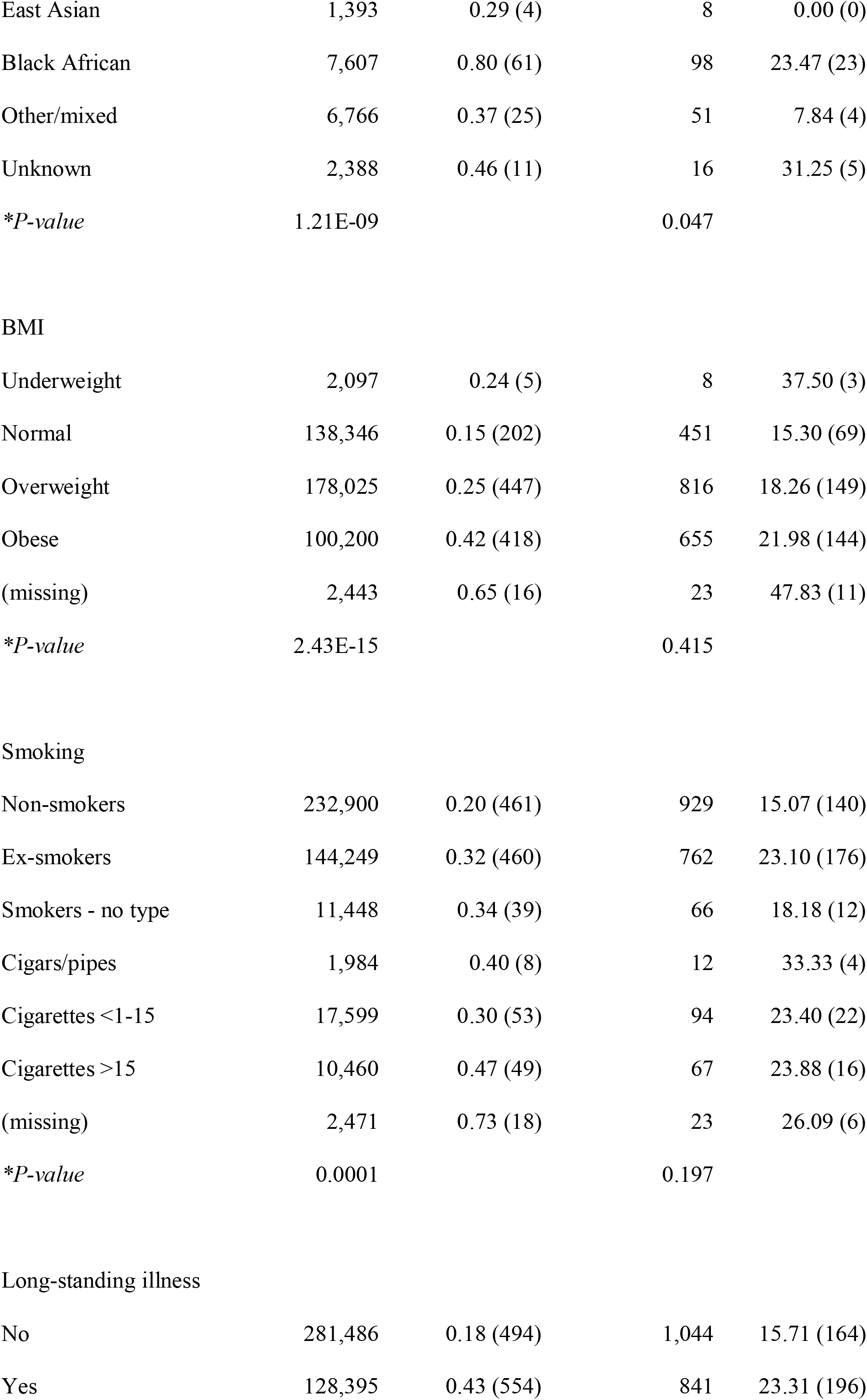
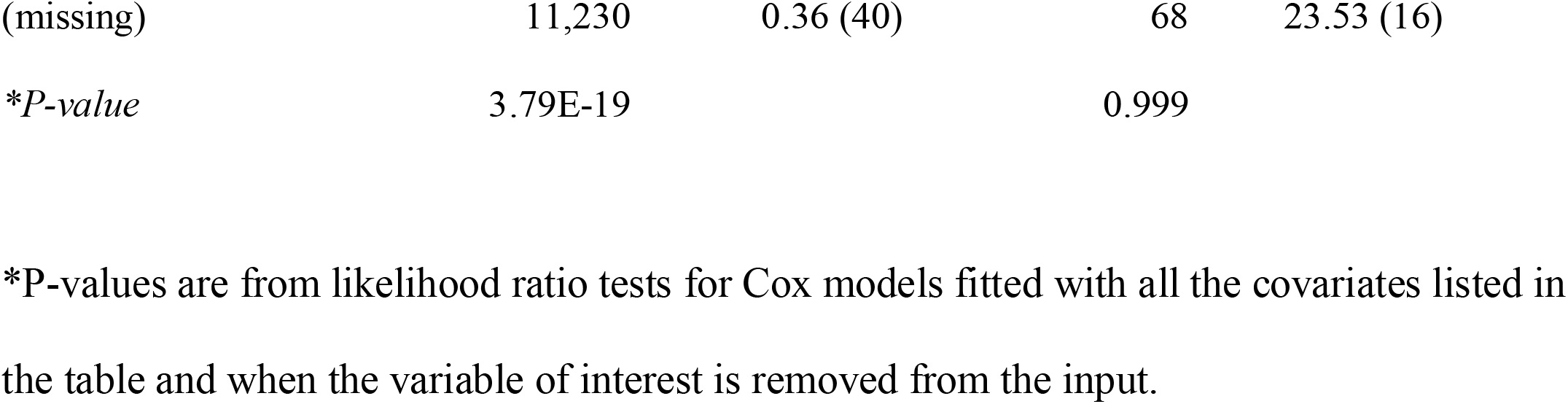
Characteristics of the UK Biobank participants in the analytical sample.

| Characteristics | Severe COVID-19 |  | COVID-19 Mortality |  |
| --- | --- | --- | --- | --- |
|  | *N* | Cases% (*N*) | *N* | Cases% (*N*) |
| Age |  |  |  |  |
| < 50 | 102,318 | 0.15 (151) | 538 | 3.35 (18) |
| 50 - 59.9 | 141,599 | 0.18 (254) | 467 | 14.56 (68) |
| 60 - 69.9 | 175,296 | 0.38 (666) | 929 | 30.68 (285) |
| 70+ | 1,898 | 0.90 (17) | 19 | 26.32 (5) |
| *\*P-value* | 4.91E-43 |  | 1.16E-34 |  |
| Sex |  |  |  |  |
| Female | 232,148 | 0.18 (419) | 921 | 14.66 (135) |
| Male | 188,963 | 0.35 (669) | 1,032 | 23.35 (241) |
| *\*P-value* | 1.12E-16 |  | 0.011 |  |
| Townsend index |  |  |  |  |
| Q1 ( -4.39, -6.26 - -3.64) | 105,211 | 0.17 (184) | 349 | 19.48 (68) |
| Q2 ( -2.92, -3.64 - -2.16) | 105,110 | 0.20 (207) | 386 | 18.13 (70) |
| Q3 ( -1.10, -2.16 - 0.48) | 105,140 | 0.25 (264) | 483 | 17.81 (86) |
| Q4 ( 2.74, 0.48 - 10.59) | 105,150 | 0.41 (432) | 734 | 20.57 (151) |
| (missing) | 500 | 0.20 (1) | 1 | 100.00 (1) |
| *\*P-value* | 0.0002 |  | 0.216 |  |
| Ethnicity |  |  |  |  |
| White European | 393,867 | 0.24 (949) | 1,698 | 19.61 (333) |
| South Asian | 9,090 | 0.42 (38) | 82 | 13.41 (11) |
| East Asian | 1,393 | 0.29 (4) | 8 | 0.00 (0) |
| Black African | 7,607 | 0.80 (61) | 98 | 23.47 (23) |
| Other/mixed | 6,766 | 0.37 (25) | 51 | 7.84 (4) |
| Unknown | 2,388 | 0.46 (11) | 16 | 31.25 (5) |
| *\*P-value* | 1.21E-09 |  | 0.047 |  |
| BMI |  |  |  |  |
| Underweight | 2,097 | 0.24 (5) | 8 | 37.50 (3) |
| Normal | 138,346 | 0.15 (202) | 451 | 15.30 (69) |
| Overweight | 178,025 | 0.25 (447) | 816 | 18.26 (149) |
| Obese | 100,200 | 0.42 (418) | 655 | 21.98 (144) |
| (missing) | 2,443 | 0.65 (16) | 23 | 47.83 (11) |
| *\*P-value* | 2.43E-15 |  | 0.415 |  |
| Smoking |  |  |  |  |
| Non-smokers | 232,900 | 0.20 (461) | 929 | 15.07 (140) |
| Ex-smokers | 144,249 | 0.32 (460) | 762 | 23.10 (176) |
| Smokers - no type | 11,448 | 0.34 (39) | 66 | 18.18 (12) |
| Cigars/pipes | 1,984 | 0.40 (8) | 12 | 33.33 (4) |
| Cigarettes <1-15 | 17,599 | 0.30 (53) | 94 | 23.40 (22) |
| Cigarettes >15 | 10,460 | 0.47 (49) | 67 | 23.88 (16) |
| (missing) | 2,471 | 0.73 (18) | 23 | 26.09 (6) |
| *\*P-value* | 0.0001 |  | 0.197 |  |
| Long-standing illness |  |  |  |  |
| No | 281,486 | 0.18 (494) | 1,044 | 15.71 (164) |
| Yes | 128,395 | 0.43 (554) | 841 | 23.31 (196) |
| (missing) | 11,230 | 0.36 (40) | 68 | 23.53 (16) |
| *\*P-value* | 3.79E-19 |  | 0.999 |  |

\*P-values are from likelihood ratio tests for Cox models fitted with all the covariates listed in the table and when the variable of interest is removed from the input.

The UK Biobank project was approved by North West Multicenter Research Ethics Committee and the National Information Governance Board for Health and Social Care (11/NW/0382). Informed consent was obtained at the time of enrolment from all participants [9]. This study was conducted under application number 20175 to the UK Biobank.

### Machine learning pipeline and Cox regression modeling

Following the pre-processing step, we conducted our analyses in two stages, namely, a) discovering predictive factors and b) epidemiological analyses as shown in Figure 1.

**Figure 1.**
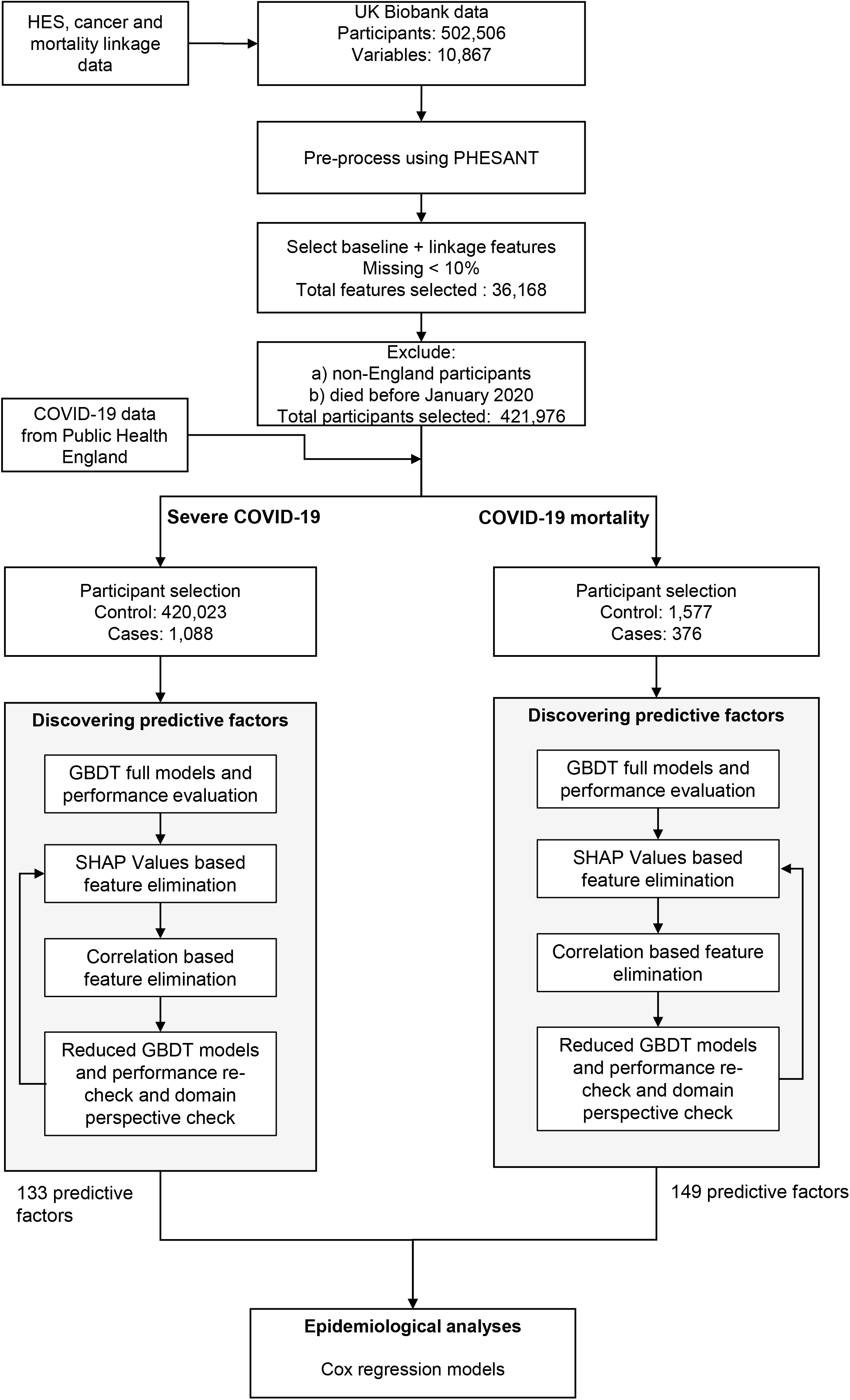
Participant selection, pre-processing, and machine learning model development pipeline for discovering predictive factors and subsequent epidemiological analyses.

#### Discovering predictive factors

Our first stage has four steps, namely, a) developing GBDT models (Supplementary Methods) with all the available features and assessing model performance, b) calculating feature importance using SHAP (Shapley Additive exPlanation) values [13, 14] and eliminating features based on a threshold, c) further elimination of highly monotonically correlated features, and d) ensuring that the reduced feature set is appropriate from a predictive performance perspective as well as from an epidemiological perspective. For our ML models, following a standard practice of internal validation in prognostic modeling, we split the severity data into random training, development, and test sets in a ratio of 60:20:20. We used the training and the development sets as the derivation cohort and the test set as the validation cohort. For mortality analysis, we split the serological samples into random training and test sets in a ratio of 80:20 due to low number of samples.

Our ML models, built using GBDT, are binary classifiers, that is, their input are the features for each individual, and their output is the model’s confidence for developing severe COVID-19 symptoms/mortality for that individual. The classes were highly imbalanced in our dataset for predicting severe COVID-19 (severe COVID-19 cases were around 0.25%) and moderately imbalanced in our dataset for predicting COVID-19 mortality (mortality rate was around 19%). To address the class imbalance problem, all our ML models were developed as weighted models [15] with the hyperparameter ‘positive class weight’ set to the ratio of negative to positive training samples, forcing GBDT to scale up gradients of the positive class samples during the training. We used CatBoost [16] (Supplementary Methods) version 0.21 implemented in Python (Python Software Foundation, version 3.5.2) for GBDT model development. GBDT model performance was assessed using the threshold independent performance metric, area under the receiver operating characteristics curve (AUROC), which has become the *de facto* standard to assess binary classifiers. AUROC confidence intervals were calculated using 1,000 bootstrap [17] datasets based on the test set for COVID-19 severity models and using 1,000 random training-test splits for COVID-19 mortality models.

For each feature, feature importance was defined as the mean absolute SHAP value as explained in Supplementary Methods. Instead of using SHAP values obtained from a single training and test cycle, we calculated SHAP values with five different randomized training and test cycles and averaged them to reduce split specific nuances. We tried a few thresholds to identify ‘important’ features and based on the features returned, we chose a threshold of 0.05% of the total importance for severity models and 0.1% for mortality models to identify ‘important’ predictive features. We used Spearman’s ρ (above 0.9) to identify sets of highly correlated features and removed all but one (the one recorded for the greatest number of samples) from those sets to produce the final set of important features to be taken to epidemiological analyses.

#### Epidemiological analyses

Following the development of univariate Cox regression models and based on the existing literature on COVID-19, we developed Cox models adjusting all models for the confounders age, sex, UK Biobank assessment center, Townsend deprivation index, ethnicity, body mass index (BMI), smoking and long-standing illness. We assessed the association of all the potential risk factors obtained from the previous stage with the outcomes, in isolation but adjusted for the confounders, for a *P*-value threshold of 0.01. We used the resulting interpretable coefficients (as opposed to mean absolute SHAP values, which do not show the directionality and are only meaningful in the context of all other features) and their 95% confidence intervals to show the association of risk in a meaningful way. We used STATA (version 16, StataCorp, College Station, TX, USA) for Cox models.

## Results

### Participants characteristics

Of the 421,111 participants included in our severity analyses, 1,088 participants (0.25%) were classified as severe COVID-19 cases based on hospital diagnoses. Of the 421,111 participants, a sub-sample of 1,953 participants, tested positive for COVID-19 or developed severe COVID-19 symptoms, and among this group there were 376 deaths and these participants were included in our mortality analyses. Table 1 shows the distribution of the participants as a whole and by categories reflecting age, sex, Townsend deprivation index, ethnicity, BMI, smoking and long-standing illness.

#### Gradient boosting decision tree (GBDT) models to discover predictors

In severity analyses, the GBDT models with all the features reported an AUROC value of 0.74 [95% CI 0.72-0.78] and the reduced features (133 features) model (after feature elimination using SHAP values and correlation) reported an AUROC value of 0.73 [95% CI 0.70-0.76] on the test set. Predictive values for mortality analyses were slightly lower, with AUROC of 0.71 [95% CI 0.68-0.73] for the full model and 0.70 [95% CI 0.65-0.74] for the reduced features (149 features) model. Supplementary Figure 1 shows the receiver operating characteristics (ROC) curves for the “all features” and “reduced features” predictive models used for SHAP value calculations. Baseline characteristics and lifestyle factors had a similar contribution to feature importance both for severity and mortality models, while sociodemographic features had a larger contribution for severity than for mortality (9.2% vs. 5.1%) (Figure 2). Health related factors including physical measures, cognitive function, self-reported disease, medications/operations, health and medical history, hospital diagnoses and biomarkers jointly accounted for > 70% of feature importance in both severity and mortality models. SHAP values for the top 50 features in severity and mortality analyses are shown in Supplementary Figure 2, with the full list including all important features in severity and mortality models shown in Supplementary Table 4 and 5 respectively. Both the severity and mortality models identified age, waist circumference and blood pressure/hypertension among the top 10 features.

**Figure 2.**
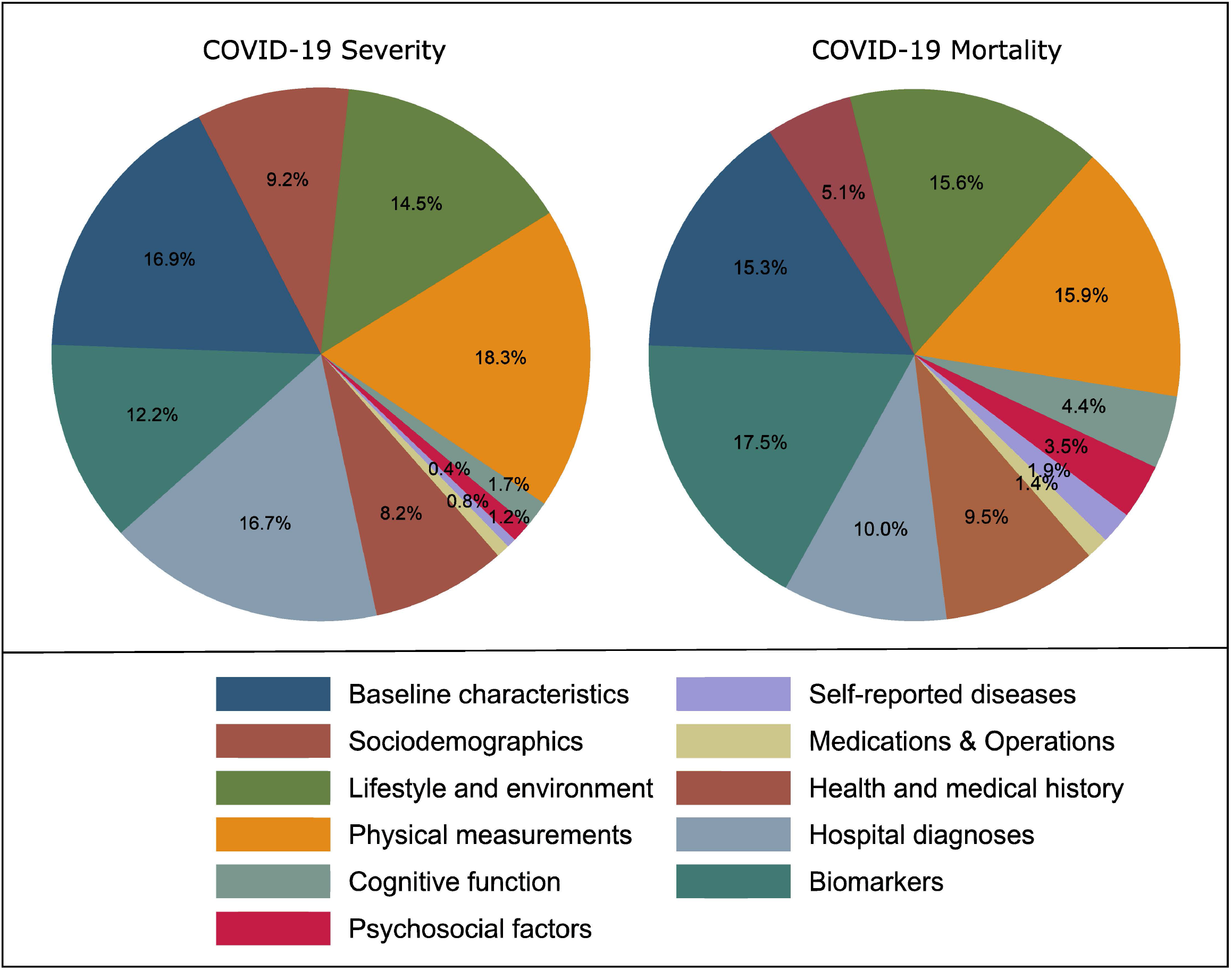
We normalize absolute mean SHAP values of all features so that the sum is equal to 100% and hence can be shown as percentages. Absolute mean SHAP feature importance values (in percentage) from reduced feature set ML models summed up category wise into eleven categories. Left: predicting severe COVID-19. Right: predicting COVID-19 mortality.

#### Epidemiological analyses

Of the 133 important features identified in the previous stage, 116 features were found to be associated with severe COVID-19 in univariate Cox models under the *P*-value threshold of 0.01, whereas 76 features were found to be associated with severe COVID-19 also in models adjusted for covariates listed in Table 1. In mortality models, of the 149 important features identified in the previous stage, 60 features were found to be associated with COVID-19 mortality in univariate Cox models for the *P*-value threshold of 0.01, whereas only 10 features were found to be associated with COVID-19 mortality after adjustment. In Supplementary Table 6 and 7 we present full data from the Cox models on severity and mortality, respectively.

Our main findings with respect to sociodemographic features are shown in Figure 3. Age had a strong association with developing severe COVID-19 symptoms with nearly 7-fold risk of severe disease in individuals >70 years vs. those < 50 years (HR 6.91, 95% CI 4.10, 11.62). Men had higher risk compared to women (HR 1.72, 95% CI 1.52-1.96) and there were clear ethnic differences; compared to white Europeans, in particular participants of black African ancestry were more likely to be affected (HR 2.79, 95% CI 2.08-3.75). Greater material deprivation (Townsend index, 4^th^ vs. 1^st^ quartile) was also associated with higher risk of severe COVID-19 disease (HR 1.43 95% CI 1.18 - 1.73). From sociodemographic features, our analyses confirmed the association between age with mortality.

**Figure 3.**
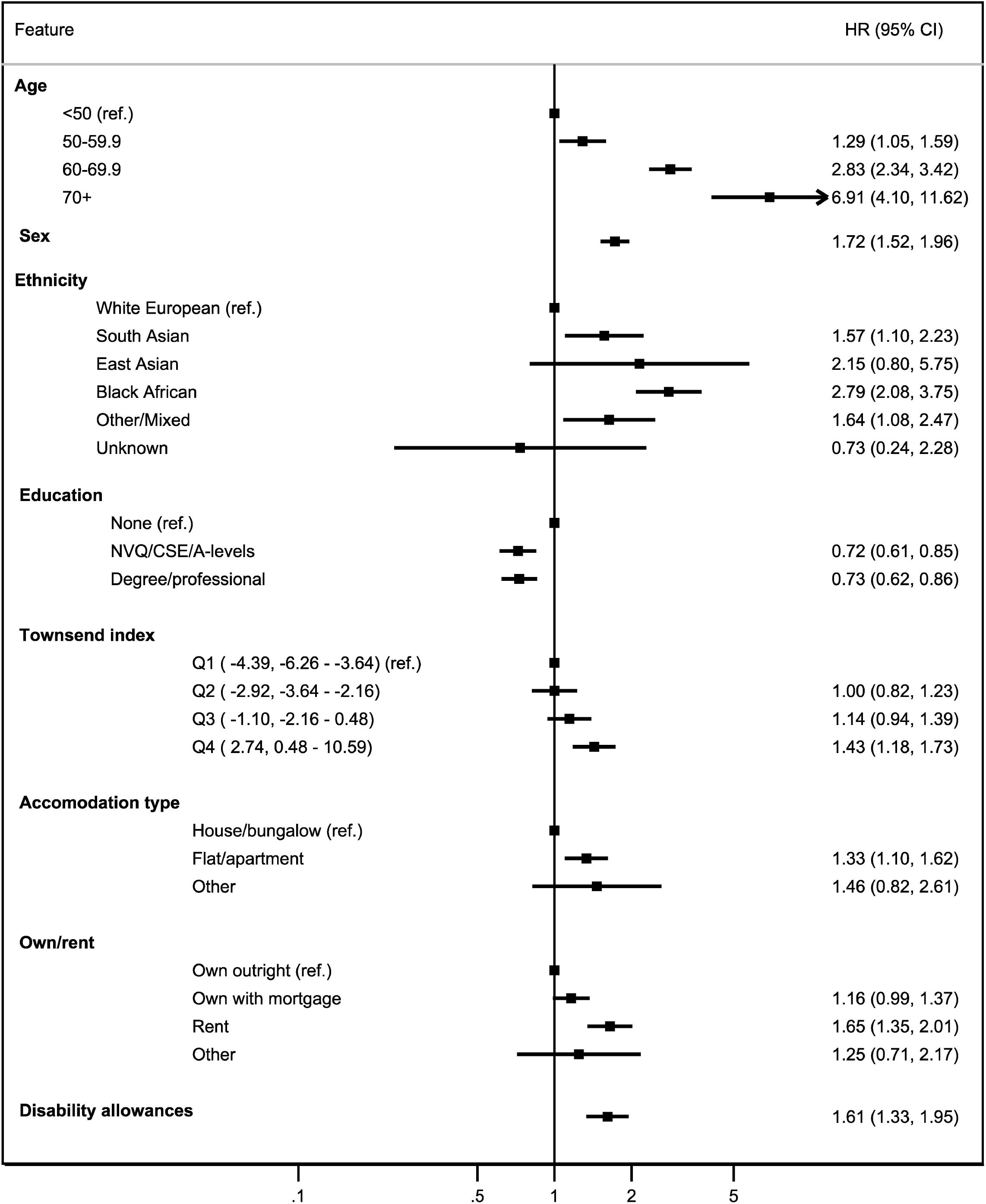
Sociodemographic factors associated with developing severe COVID-19 symptoms with their hazard ratios and 95% confidence intervals from Cox regression models adjusted for age, sex, UK Biobank assessment center, Townsend deprivation index, ethnicity, body mass index (BMI), smoking, and long-standing illness.

As shown in Figure 4 describing associations identified for lifestyle factors, compared to non-smokers, ex-smokers and current smokers had a higher risk of developing severe COVID-19. Those who smoked more than 15 cigarettes a day were found to be associated with the highest risk in the smoking categories (HR 2.25 95% CI1.34 - 3.78). Our results show that, in general, physical activity was associated with decreased risk, while greater inactivity reflected by more time spent watching television is associated with increased risk. We also found tea intake to be associated with decreased risk of developing severe COVID-19 symptoms. We did not find strong evidence for associations between baseline lifestyle factors and mortality risk, except that for current smoking (smoked on all or most days), which was associated with increased risk of COVID-19 mortality (HR 1.41 95% CI 1.01 – 1.99).

**Figure 4.**
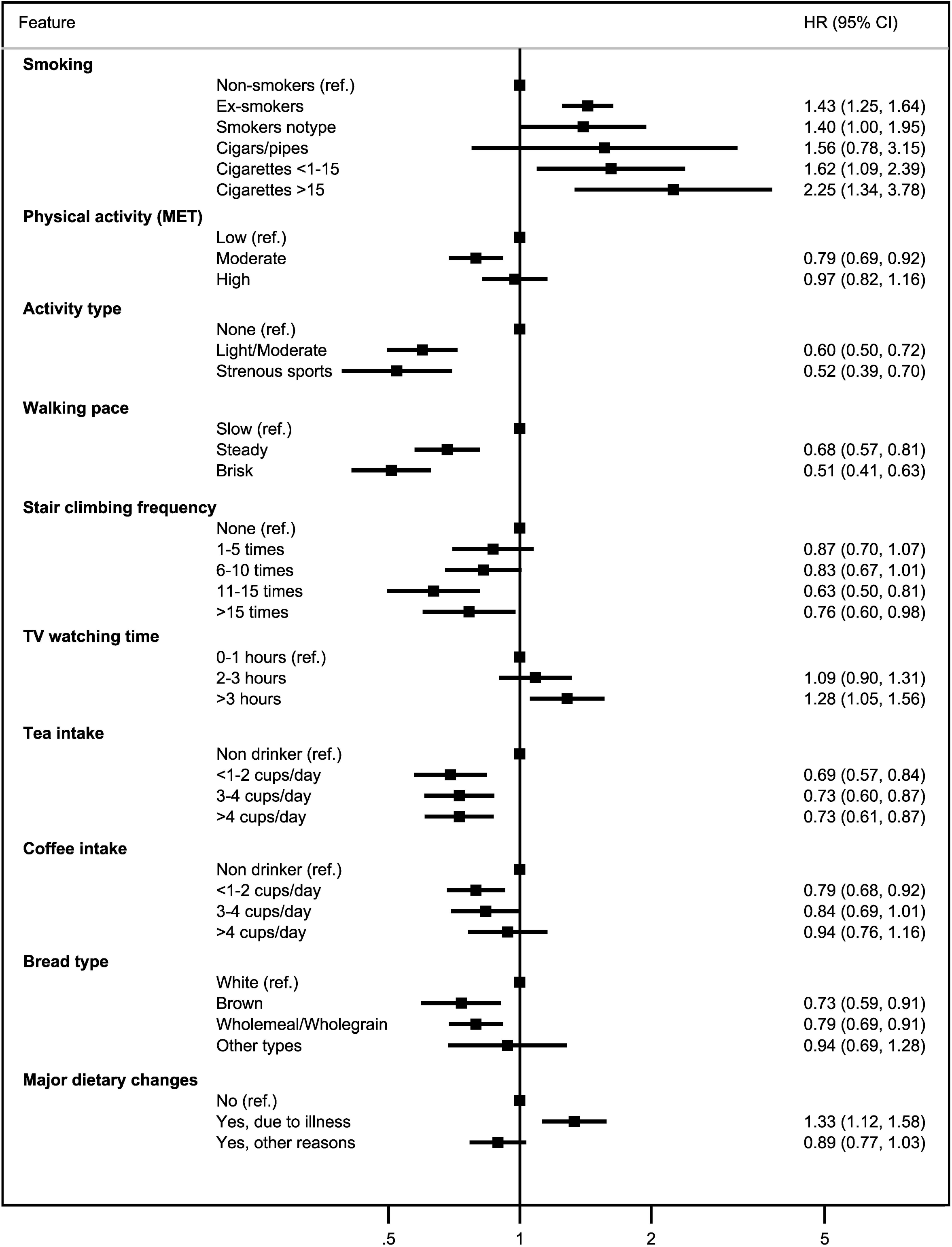
Lifestyle factors associated with developing severe COVID-19 symptoms with their hazard ratios and 95% confidence intervals from Cox regression models adjusted for age, sex, UK Biobank assessment center, Townsend deprivation index, ethnicity, body mass index (BMI), smoking, and long-standing illness.

Figure 5 describes associations between health-related outcomes and severe COVID-19 diseases. Worse self-rated health status and higher number of treatments/medications were associated with developing severe COVID-19 symptoms. From the disease outcomes, prior diagnoses of lung disease (including wheezing, pneumonia, and COPD), type 2 diabetes, hypertension, and urinary system disorders were all associated with an increased risk. Further associations were seen with higher risk of severe disease by anemia (HR 2.24 95% CI 1.80 – 2.78), nausea and vomiting (HR 2.15 95% CI1.70 - 2.70), depression (HR 2.12 95% CI1.70 - 2.65) and psychoactive substance abuse (HR 1.67 95% CI 1.41 - 1.97). From serum biomarkers measured at the baseline (i.e., 10 to 14 years before COVID-19 diagnoses), we found evidence for an adverse association by higher levels of Cystatin C, C-reactive protein (CRP), gamma glutamyl transferase (GGT), and alkaline phosphatase (ALP), in addition to a higher risk of developing severe COVID-19 by microalbuminuria (Figure 6). In mortality analyses, prior primary diagnosis of urinary tract infection was associated with COVID-19 mortality (HR 1.95 95% CI 1.30 – 2.91). We did not find strong evidence for associations between baseline serum biomarkers and COVID-19 mortality.

**Figure 5.**
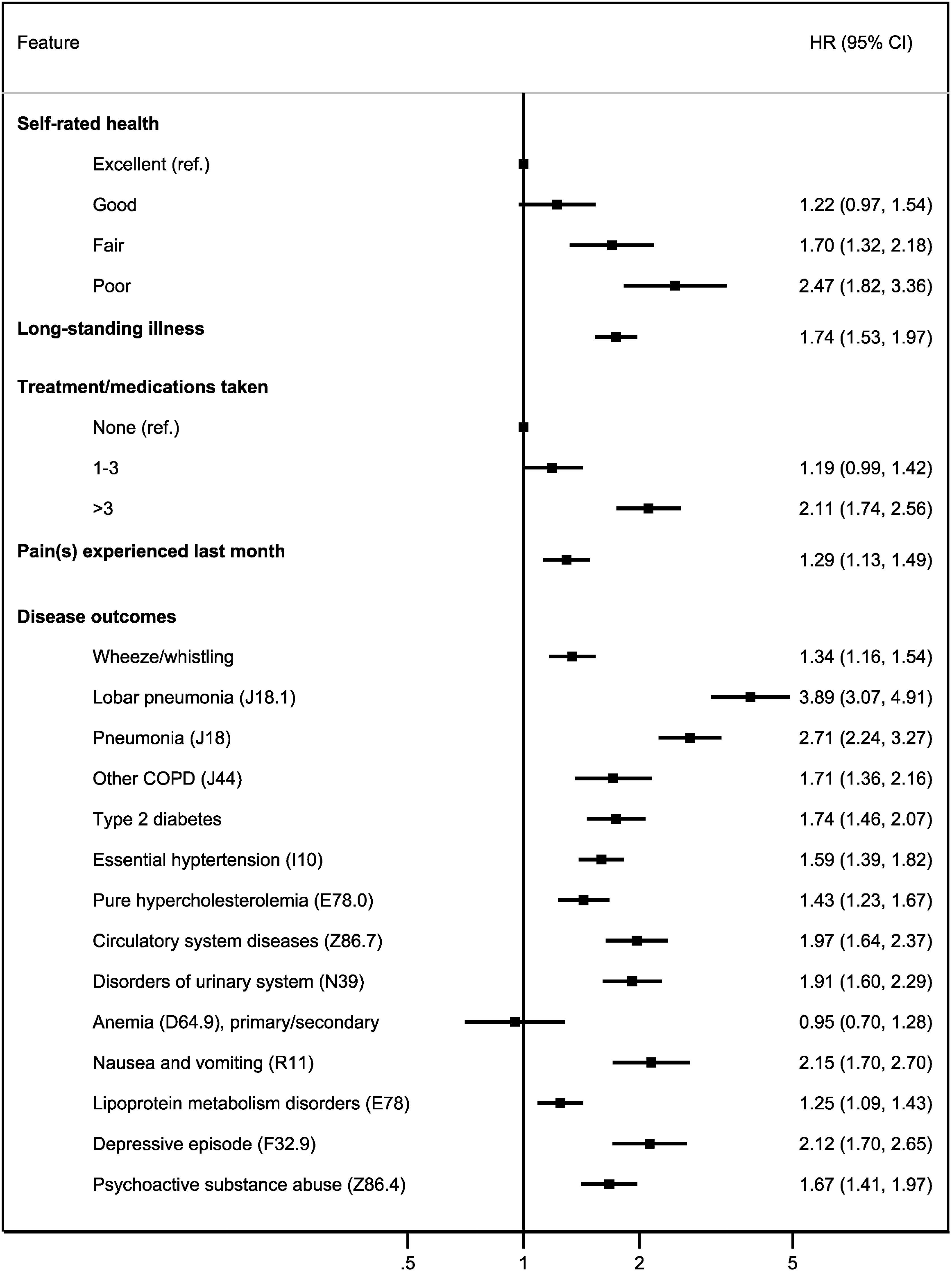
Self-rated health, medications, and disease outcomes associated with developing severe COVID-19 symptoms with their hazard ratios and 95% confidence intervals from Cox regression models adjusted for age, sex, UK Biobank assessment center, Townsend deprivation index, ethnicity, body mass index (BMI), smoking, and long-standing illness.

**Figure 6.**
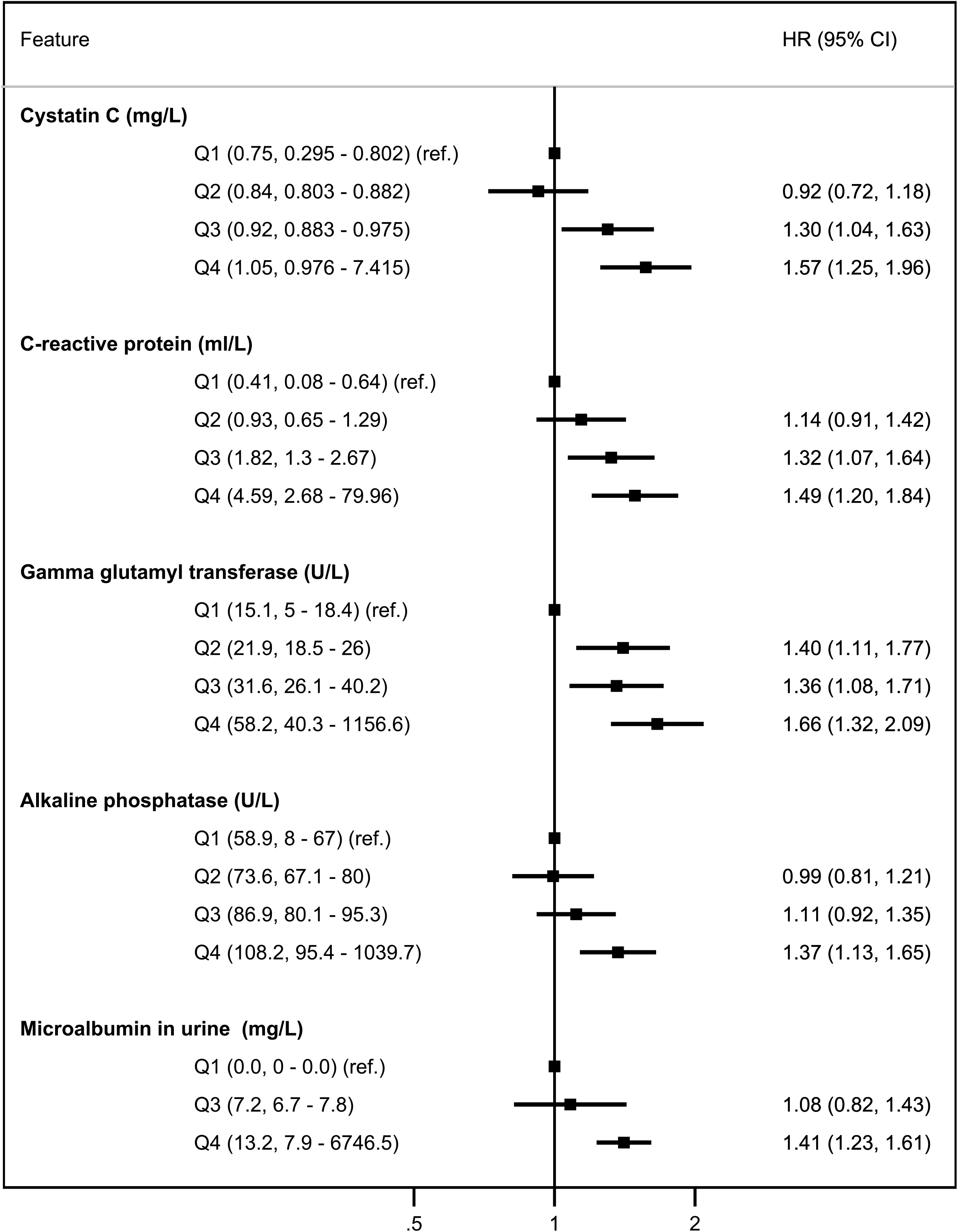
Baseline biomarkers associated with developing severe COVID-19 symptoms with their hazard ratios and 95% confidence intervals from Cox regression models adjusted for age, sex, UK Biobank assessment center, Townsend deprivation index, ethnicity, body mass index (BMI), smoking, and long-standing illness.

## Discussion

The purpose of this study was to identify characteristics reflecting longer-term susceptibility to infection, in particular, susceptibility to severe COVID-19 disease and mortality. We used a novel approach of combining ML with conventional epidemiological analyses, which allowed us to identify a number of possible risk factors from a very large pool of features when assessing associations with COVID-19 risk. While our analyses confirmed associations for many well-known population determinants and risk factors for COVID-19 disease, our study also suggested possible predictive roles for renal and liver blood biomarkers as reflecting longer-term susceptibility to infection. Our results highlighted the increased risks for those members of society who are the most deprived, while associations observed for lifestyle factors largely supported current general advice for better general health.

Pre-existing disease, in particular respiratory conditions, type 2 diabetes and hypertension and other cardiovascular diseases, were expectedly associated with severe COVID-19 disease. Our study shows similar results obtained by studies conducted elsewhere (i.e., not in England) on COVID-19 severity and mortality. For example, in line with previous studies [18-23], our study also shows evidence of association between comorbidities, hypertension and diabetes and developing severe COVID-19 symptoms, although such associations were less apparent in our mortality analyses.

Both depression and psychoactive substance abuse were associated with increased risk of severe COVID-19 disease and the magnitude of this effect was similar compared to that observed for purely physiological diseases. In line with our earlier phenome-wide investigation which suggested wide ranging downstream health effects for depression [24], these findings highlight the need to monitor and treat physical diseases in those affected.

Consistent with previous studies, our study showed the disproportional number of people who developed severe COVID-19 symptoms from black African or ‘other’ (unspecified) ethnic backgrounds [25]. Also, in line with previous studies, our study picked up disparities in severe COVID-19 cases with respect to multiple aspects of social and material deprivation, including lower education, type of accommodation, and home ownership, highlighting the need to consider infection control as an important aspect when addressing inequalities in health. In line with a previous study looking at the determinants of a positive test result in the UK Biobank [26], we confirmed the key predictors including ethnicity, male sex and higher BMI were associated with testing positive for COVID-19 and our study shows such an association exists also for developing severe symptoms. We also observe similar associations with respect to lower education attainment and testing positive [27] and developing severe COVID-19 symptoms.

We found that higher levels of indicators of renal dysfunction (cystatin C and urinary microalbumin) and disorders of the urinary system were associated with severe COVID-19 disease. Furthermore, biomarkers of liver injury (GGT and ALP) and systemic inflammation (CRP) were also associated with severity. Indicators of acute renal dysfunction, elevated liver metabolism and inflammation are frequently reported to accompany the disease course of COVID-19, and to associate with more severe COVID-19 outcomes [28-32]. Respiratory diseases including pneumonia and COPD are known to increase the risks related to reduced oxygen carrying capacity in COVID-19, and hypoxemia in turn has been identified as a factor affecting liver function in critical COVID-19 cases [33]. However, in this study biomarkers reflecting renal dysfunction and liver injury were in most cases measured more than a decade earlier, suggesting that these biomarkers *per se* may reflect a long-term susceptibility to severe infection.

A number of studies have used the UK Biobank to investigate risk factors associated with developing severe COVID-19 symptoms and COVID-19 mortality. The uniqueness of our study can be attributed to more than one aspect. Many studies focus on one or two particular risk factors (e.g., [34, 35]) or an area of interest such as sociodemographic (e.g., [36]), lifestyle factors (e.g., [37]), disease outcomes, and biomarkers (e.g., [23, 38]), and choose the predictors in advance. Our novel hypothesis-free approach of combining ML and traditional epidemiological methods, investigates all predictors (such as sociodemographic, lifestyle, and psychosocial factors, cognitive functions, biomarkers, disease outcomes and treatments). While we reported associations for the well-known risk factors, our analyses also suggest long-term predictive ability for markers of liver and kidney injury, and inflammation.

There are some methodological considerations which need to be considered in the context of our study. We defined severe COVID-19 based on hospital episode statistics and looked at predictors of mortality only in the subgroup with a positive test/known infection. Other studies have defined COVID-19 infection based on a positive test result from RT-PRC tests [38, 39] in the subgroup tested for infection, while testing positive for COVID-19 within a particular time period (when most of the testing took place in a hospital setting) has been used as a proxy for COVID-19 severity [40, 41]. While our outcome will only capture COVID-19 cases which required hospitalization, it allows us to assess long-term predictors of severe infection using information from the whole cohort. Some previous studies have looked into all-cause mortality subsequent to testing positive for COVID-19 mortality [42], while our approach was more specific and required cause of death mentioning COVID-19 related disease codes (U07.1 or U07.2).

While the UK Biobank prospective cohort is unique in its size and scope with extensively phenotyped and genotyped data, enabling hypothesis-free approaches for identifying long-term predictors of infection risk, it is also a cohort of volunteers with higher education and socio-economic status, and lower mortality rates compared to the general population [43]. The healthy volunteer bias may have affected our analysis and thus the external validity. However, it was reassuring that several of our findings have been observed in studies conducted in other parts of the world. Our mortality analyses were conducted in the sub-sample testing positive for COVID-19, hence, we may have lacked power to detect further predictors. Also, while our results show association and not causation, we are unable to discount residual confounding by factors not included in our analyses.

In conclusion, our large-scale hypothesis-free approach identified several risk factors associated with COVID-19 infection, and suggested indicators of renal dysfunction, liver injury and inflammation as predictors of long-term infection risk. Our data also highlights the need to focus on infection control in attempts to reduce inequalities in health.

## Data Availability

Data can be accessed through the UK Biobank.

