## supplemental materials for "Identifying risk factors for COVID-19 severity and mortality in the UK Biobank"

**Supplementary Methods**

**Background of GBDT**

Gradient boosting decision trees (GBDT) are ensembles of decision trees that are constructed in a sequence such that each subsequent tree after the first tree is trained to predict the error between the observed and predicted value obtained to that point. They are trained using supervised learning. Each decision tree is built by splitting the entire training samples into smaller and smaller groups successively until a predefined condition such as depth of the tree (as determined using a hyper-parameter) or other termination conditions are met. Each training sample will belong to only one leaf of a tree and, in our application, each leaf predicts some level of COVID-19 severity/mortality. Features and their values at which the split is to be made to create two new branches to successively build a tree, are chosen by the algorithm in order to optimize an objective function. A development set can be used to avoid overfitting by limiting the number of such decision trees created. GBDTs are currently considered to be the state-of-the-art supervised learning algorithm for building predictive models using tabular data. A study conducted to assess the performance of 13 state-of-the-art machine learning algorithms on a set of 165 publicly available classification problems (mostly bioinformatics problems) found GBDT to be the top ranked in mean ranking across the problems.[1] There are end-to-end implementations of GBDTs, capable of handling billions of samples and millions of variables such as XGBoost [2], LightGBM [3] and CatBoost [4]. They utilize graphical processing units (GPUs) in addition to central processing units to improve training and inference speed. Each machine learning algorithm has its own hyperparameters, also known as tuning parameters and that can control the behavior of an algorithm. Examples of such hyperparameters include learning rate, depth of decision trees, maximum number of decision trees and so on. Generally, CatBooost outperforms XGBoost and LightGBM in performance using default hyperparameters.[5, 6] Also, improvements in performance using tuned hyperparameters may not be largely different from performance with default hyperparameters as seen in some studies using CatBoost.[6] CatBoost provides native support to handle categorical features (with or without numeric values). It can be instructed to consider missing value as an instance of value, guaranteeing a split between missing value and other non-missing values while trees are built.

**Specific GBDT methods**

For CatBoost, the most important hyperparameters are learning rate and number of trees to be built (also known as number of iterations or number of estimators). If used with default values, CatBoost dynamically selects learning rate based on number of iterations (default value of iterations is 1,000). For our experiments for predicting COVID-19 severity we set the number of iterations to 2,000 and used a development set to avoid overfitting by stopping growing new trees when there is no improvement in area under the receiver operating characteristics curve (AUROC) performance on the development set in 50 consecutive iterations. For our COVID-19 mortality predictions, we set number of iterations to its default value. Models were set to utilize GPUs for predicting COVID-19 severity. Boosting type was set to plain boosting. All other hyper-parameters were left at their default values. We used features without imputing missing values and hospital diagnoses.

**Pre-processing using PHESANT**

PHESANT [7] classifies variables as continuous, ordinal, and categorical using a rule-based system to determine the appropriate coding of each variable. It also deals with various scenarios such as handling of multiple initial measurement of variables (e.g. spirometry), coding unusual values (e.g. negative values are used to code answers such as ‘Preferred not to answer’ and ‘Don’t know’) as missing, changing the order of variable values to make them logical (e.g., field 1239, current smoking status) and creating proper dummy variables (e.g., secondary diagnoses field 41204 had 184 array elements and any ICD10 code could be stored in any array elements).

**SHAP (Shapley Additive explanation) Values**

SHAP values are based on Shapley values (derived by Lloyd Shapley in 1953 [8]), a solution concept in game theory. Shapley values deal with how fairly (by satisfying certain conditions) the payoffs from a cooperative game can be distributed to game players. Shapley value for a player is defined as the average marginal contribution of that player, considering all possible combinations that the player can be part of.

SHAP values is a local additive feature (features and predictors are used here synonymously) attribution method [9], i.e., it contrastively explains each observation in isolation using a linear function with predicted output as function output and simplified and interpretable feature values as input. Since SHAP values use a linear approximation, for binary classifiers transforming margins using logistic function, SHAP values will be in log-odds space. SHAP values are consistent with respect to feature attribution measurement in tree models, even in the presence of correlated features. If *f* represents the ML model learned (e.g., a trained GBDT model) and *g* represents the local linear explanation model (for a particular observation, $\boldsymbol{x}$), then

$f\left( \boldsymbol{x} \right)=g\left( \boldsymbol{x}^{'} \right)= \emptyset_{0}+ \sum_{i=1}^{M} \emptyset_{i}x_{i}^{'}$,

where, $\emptyset_{0}$ is the output when no input is present, *M* is the number of features, $\boldsymbol{x}^{\boldsymbol{'}}$ is the simplified, dichotomized and interpretable vector representing ***x*** in the local explanation model and $\emptyset_{i}\mathbb{\in R}$ is the attribution to each feature. According to [9], $\emptyset_{i}$ given *f* and ***x*** and satisfying some desirable conditions (local consistency and missingness) is given as [9]

$$\emptyset_{i}\left( f,\boldsymbol{x} \right)= \sum_{\boldsymbol{z}^{\boldsymbol{'}}\subseteq\boldsymbol{x}^{'}} \frac{\left| \boldsymbol{z}^{'} \right|!\left( M-\left| \boldsymbol{z}^{'} \right|-1 \right)!}{M!} \left[ f_{\boldsymbol{x}}\left( \boldsymbol{z}^{'} \right)- f_{\boldsymbol{x}}\left( \boldsymbol{z}^{'}\backslash i \right) \right],$$

where, $\left| \boldsymbol{z}^{'} \right|$ is the number of features present (non-zero elements) in $\boldsymbol{x}^{\boldsymbol{'}}$. The solution to the above equation is the Shapley values of a conditional expectation function of the original model *f*. We use the recent implementation, called TreeSHAP [10] specifically developed for tree-based models such as random forest and gradient boosting. TreeSHAP can calculate SHAP values much faster by reducing computational complexity from $\mathcal{O}\left( TL2^{M} \right)$ to $\mathcal{O}\left( TLD^{2} \right)$ (*T* is the number of trees, *L* is the number of leaves, *M* is the number of features, and *D* is the depth of the tree) compared to the previous implementation KernalSHAP [9]. We used the Python package ‘SHAP’ version 0.34 for calculating SHAP values.

**Feature selection using SHAP values and correlation**

For each feature, we calculated feature importance as the mean absolute SHAP value in the training set, as

$\varphi_{i}= \frac{1}{N} \sum_{j=1}^{N} |\emptyset_{ji}|$,

where *N* is the total number of observations in the training set.

We normalized mean absolute SHAP values ($\varphi_{i})$ to 100% and a cut-off value of 0.05% was used to identify ‘important’ features for severe COVID-19 and 0.1% for COVID-19 mortality. We used Spearman’s ρ (above 0.9) to identify sets of highly correlated features and removed all but one (the one recorded for the greatest number of samples) from those sets to produce the final set of important features. We plotted SHAP values of important features for each sample to understand the direction and magnitude of impact of individual features on model output.
