## supplementary figure 1 for "Identifying risk factors for COVID-19 severity and mortality in the UK Biobank"

| 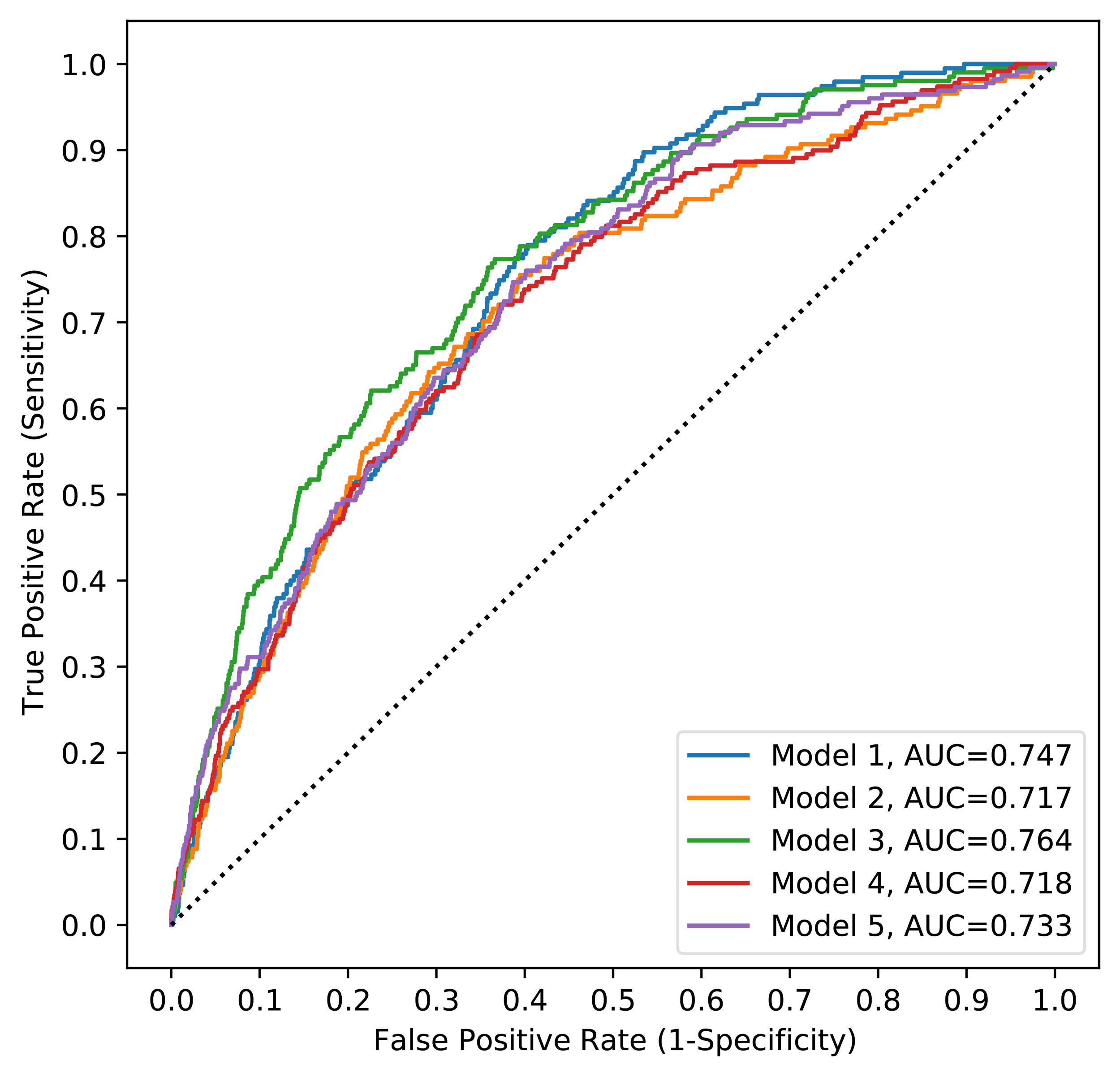 | 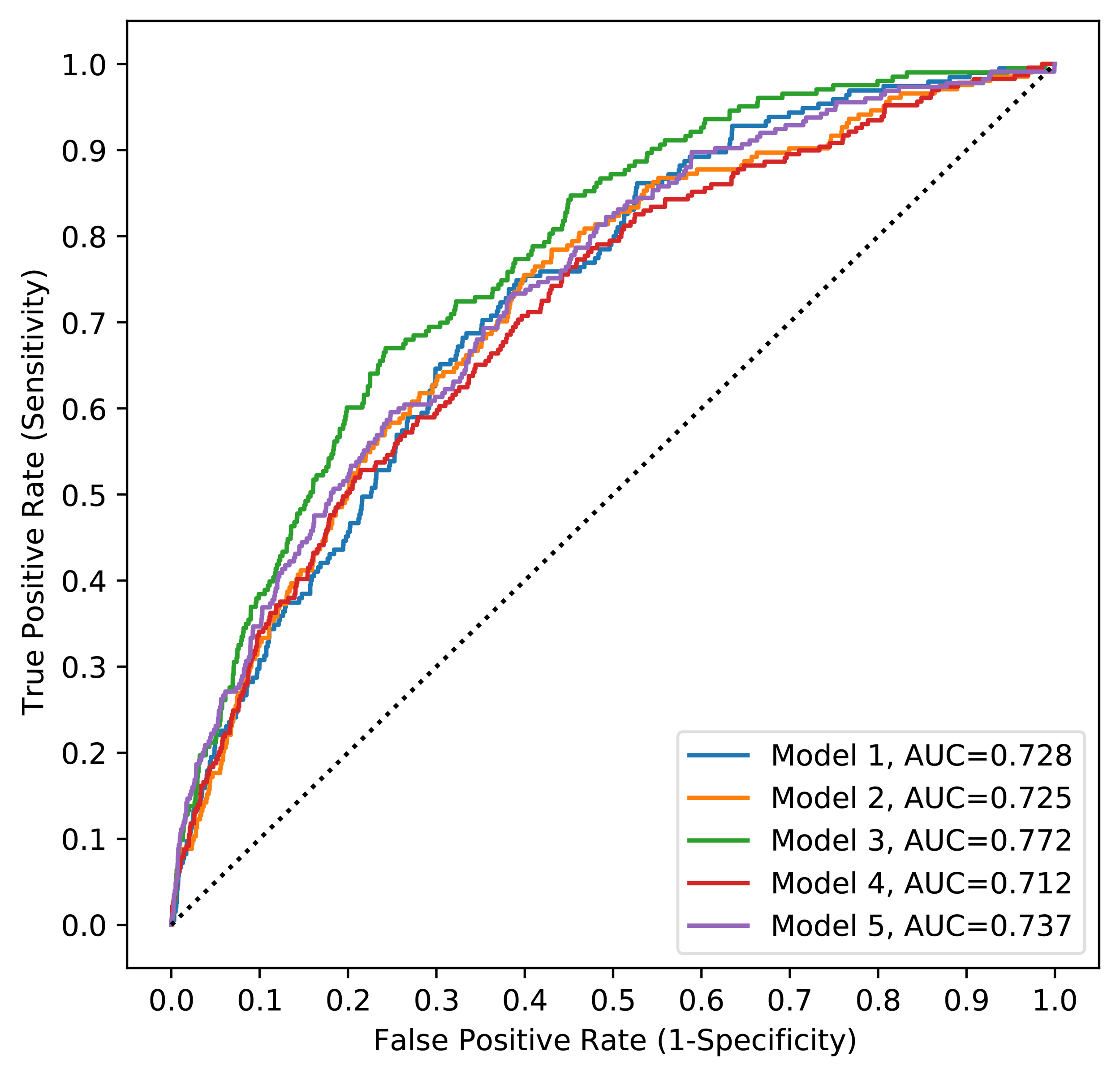 |
| --- | --- |
| 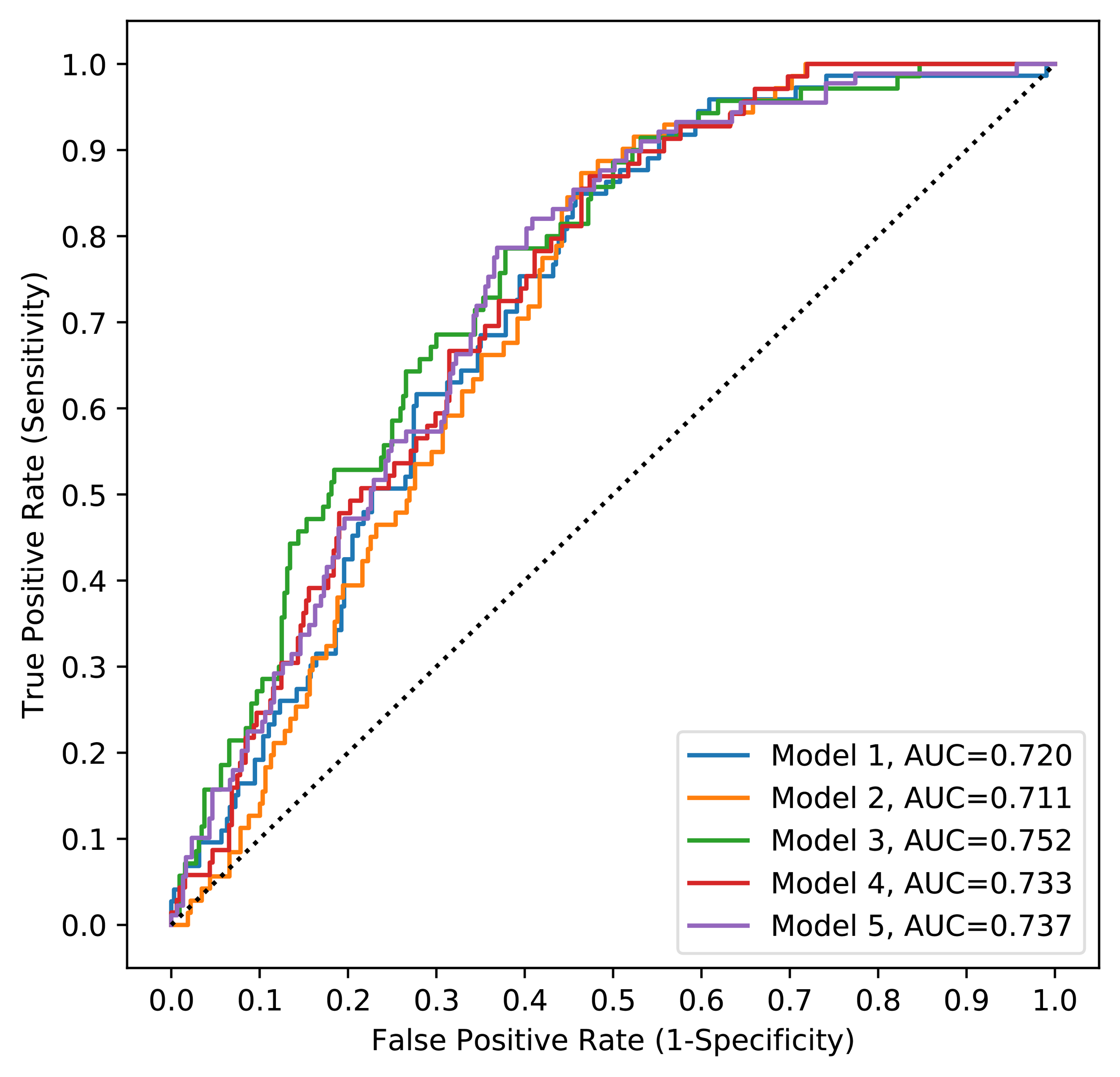 | 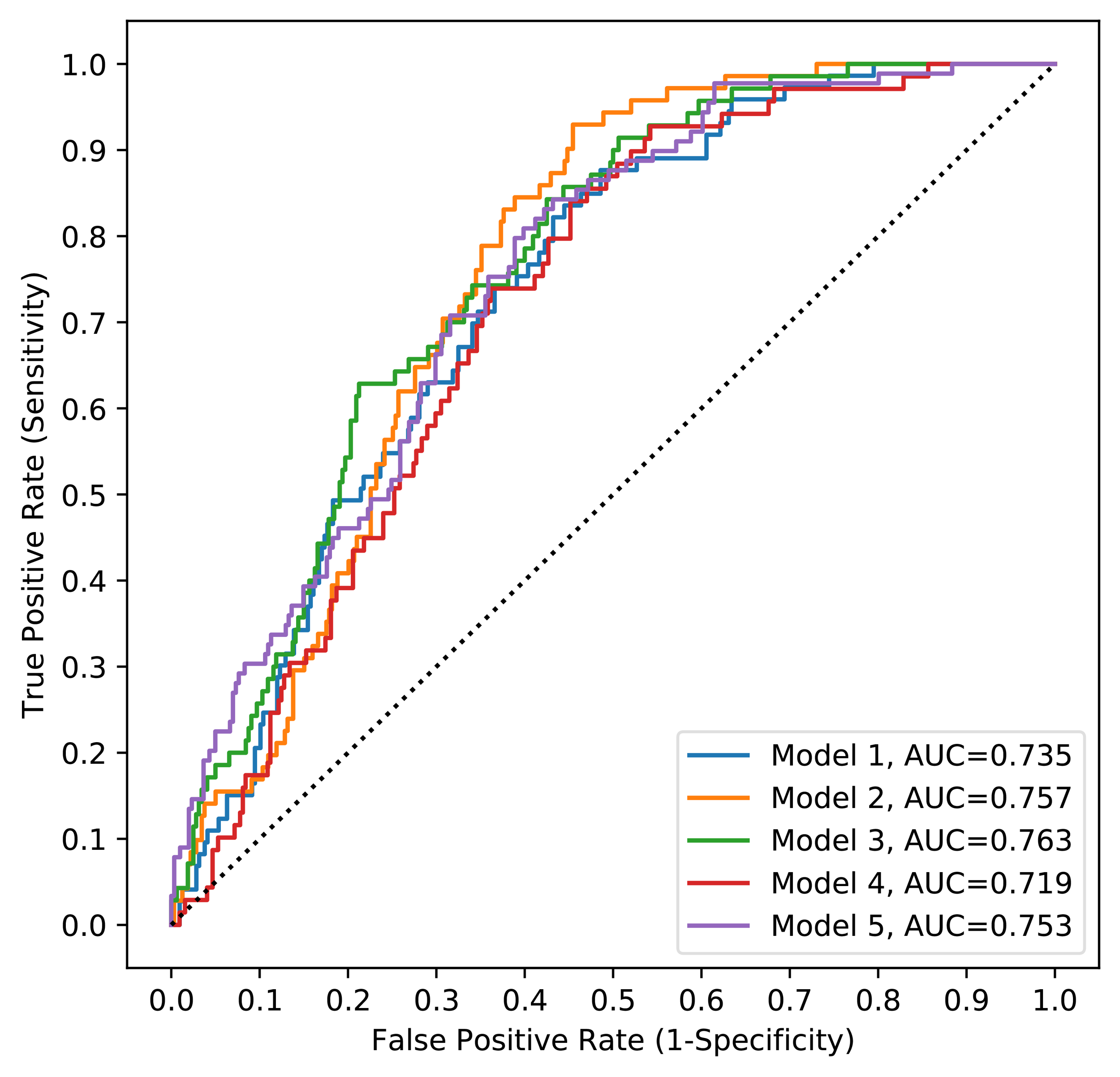 |

**Supplementary Figure 1**. In each scenario explained, five models by developing models based on splitting the data randomly five times. ***Upper left***: receiver operating characteristics (ROC) curves of five gradient boosting decision trees (GBDT) COVID-19 severity models when all the features were used for calculating average SHAP values across the models. ***Upper right***: receiver operating characteristics (ROC) curves of five gradient boosting decision trees (GBDT) COVID-19 severity models when only the important features (133 features) used for calculating average SHAP values across the models. ***Lower left***: receiver operating characteristics (ROC) curves of five gradient boosting decision trees (GBDT) COVID-19 mortality models when all the features were used for calculating average SHAP values across the models. ***Lower right***: receiver operating characteristics (ROC) curves of five gradient boosting decision trees (GBDT) COVID-19 mortality models when only the important features (149 features) used for calculating average SHAP values across the models.
