## supplementary figure 2 for "Identifying risk factors for COVID-19 severity and mortality in the UK Biobank"

| A)  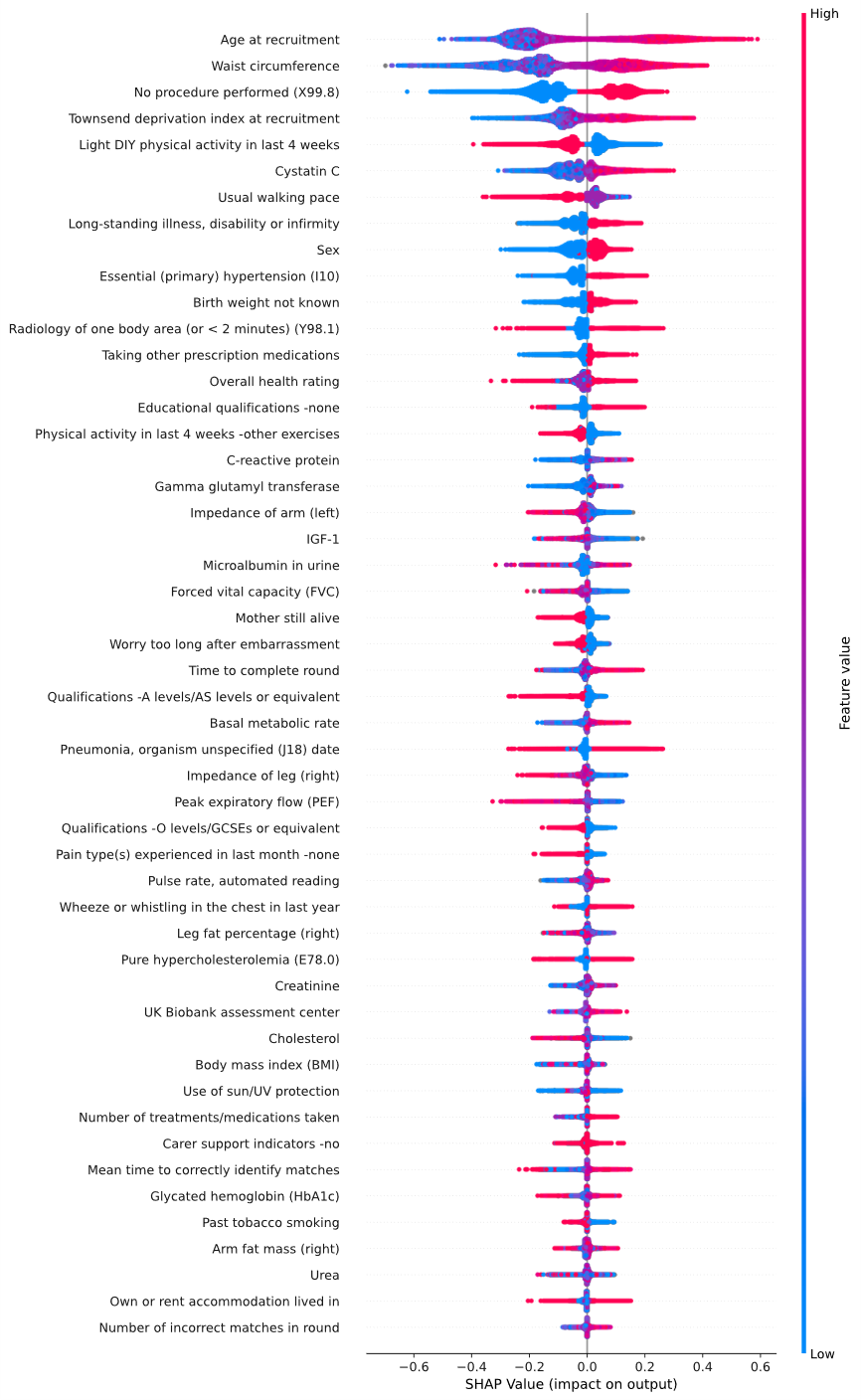 | B)  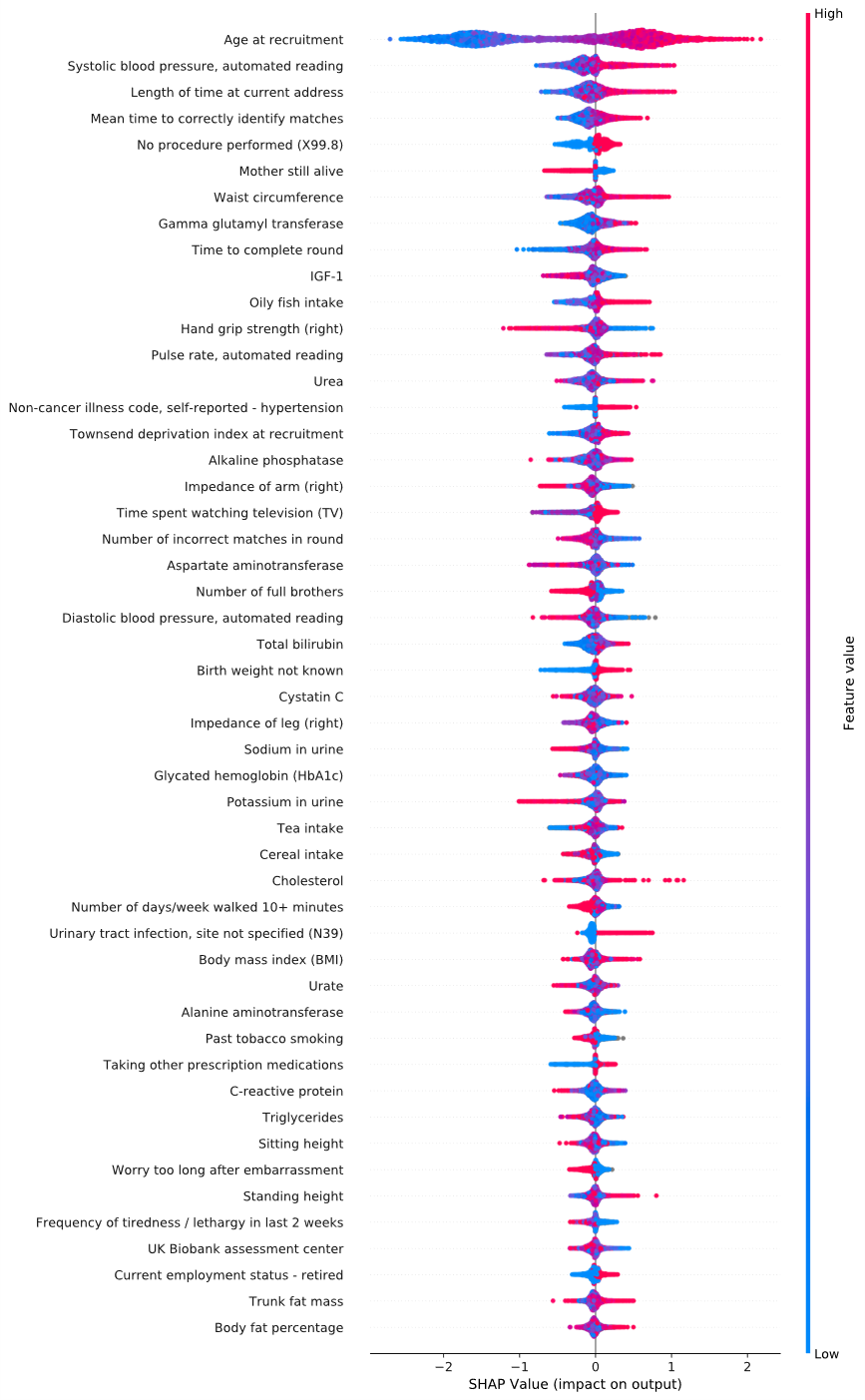 |
| --- | --- |

**Supplementary Figure 2**. SHAP summary plot showing the top 50 features from GBDT models using reduced number of features for predicting severe COVID-19 (in panel A) and for predicting COVID-19 mortality (in panel B). Each point on the plot is a SHAP value for a feature and a sample. The position on the y-axis is determined by the feature and on the x-axis by the SHAP value. The color gradient represents the feature values from low (blue) to high (red). Grey dots represent samples with missing values for a feature.
