## supplementary table 3 for "Identifying risk factors for COVID-19 severity and mortality in the UK Biobank"

**Supplementary Table 3**. Category wise count of features before and after pre-processing using PHESANT for predicting severe COVID-19 and predicting COVID-19 mortality. Counts of features for mortality prediction is given in brackets if they differ from the counts of features for severe COVID-19 prediction.

| **Category** | **All features** | | **Important features** | | **Important features** | |
| --- | --- | --- | --- | --- | --- | --- |
|  |  |  |  |  | **(after dropping correlated features)** | |
|  | **UK Biobank fields** | **Derived features** | **UK Biobank fields** | **Derived features** | **UK Biobank fields** | **Derived features** |
| A - Baseline characteristics | 6 | 6 | 6 | 6 | 4 | 4 |
| B - Sociodemographics | 12 | 38 | 7 | 10 (8) | 7 | 10 (7) |
| C - Lifestyle and environment | 52 (50) | 65 (63) | 26 (30) | 29 (31) | 26 (30) | 29 (31) |
| D - Physical measurements | 52 (45) | 52 (45) | 44 (43) | 44 (43) | 17 | 17 |
| E - Cognitive function | 6 | 6 | 4 | 4 | 3 | 3 |
| F - Psychosocial factors | 20 | 26 | 5 (10) | 6 (10) | 5 (10) | 6 (10) |
| G - Self-reported diseases | 4 | 511 | 2 | 2 (4) | 2 | 2 (4) |
| H - Medications and Operations | 5 | 2029 | 4 (3) | 5 (6) | 4 (3) | 5 (6) |
| I - Health and medical history | 30 (29) | 96 (83) | 17 | 18 (22) | 16 (17) | 17 (21) |
| J - Hospital diagnoses | 1,031 | 33,291 | 14 (17) | 30 (35) | 11 (15) | 22 (27) |
| K – Biomarkers | 48 (47) | 48 (47) | 20 | 20 | 18 | 18 |
| **Total** | **1,266 (1,255)** | **36,168 (36,145)** | **149 (159)** | **174 (189)** | **113 (126)** | **133 (149)** |
