## supplementary table 4 for "Identifying risk factors for COVID-19 severity and mortality in the UK Biobank"

**Supplementary Table 4**. Listing of the important features identified using SHAP values for predicting severe COVID-19. Three sets of SHAP values are provided: a) SHAP values from GBDT models with all the features in their input totalling 73%, b) SHAP values from GBDT models using only the important features (SHAP value >= 0.05% when all the features were input), and c) SHAP values from GBDT models with only the important features in their input, after removing 41 highly correlated features (correlation above 0.9).

| **Feature ID** | **Description** | **SHAP value (all features)** | **SHAP value (important features)** | **SHAP value (after removing highly correlated features)** |
| --- | --- | --- | --- | --- |
| x21022 | Age at recruitment | 3.52 | 5.36 | 13.58 |
| x48 | Waist circumference | 8.91 | 10.65 | 10.43 |
| x41272__X998 | No procedure performed (X99.8) | 3.83 | 6.37 | 8.66 |
| x189 | Townsend deprivation index at recruitment | 5.24 | 5.52 | 5.34 |
| x6164__4 | Light DIY physical activity in last 4 weeks | 3.50 | 4.23 | 3.91 |
| x30720 | Cystatin C | 3.39 | 3.57 | 3.73 |
| x924 | Usual walking pace | 3.36 | 3.35 | 2.81 |
| x2188 | Long-standing illness, disability or infirmity | 1.83 | 2.14 | 2.61 |
| x31 | Sex | 1.79 | 1.88 | 2.49 |
| x41204__I10 | Essential (primary) hypertension (I10) | 1.57 | 1.50 | 2.44 |
| x120 | Birth weight known | 1.83 | 1.97 | 2.34 |
| x41272__Y981 | Radiology of one body area (or < 2 minutes) (Y98.1) | 0.65 | 0.93 | 1.39 |
| x2178 | Overall health rating | 1.61 | 1.88 | 1.37 |
| x6138__100 | Educational qualifications - none | 1.36 | 1.19 | 1.28 |
| x6164__2 | Physical activity in last 4 weeks - other exercises | 0.81 | 0.88 | 1.28 |
| x2492 | Taking other prescription medications | 1.39 | 1.26 | 1.23 |
| x23110 | Impedance of arm (left) | 0.33 | 0.34 | 1.13 |
| x30710 | C-reactive protein | 0.71 | 0.44 | 1.12 |
| x30730 | Gamma glutamyl transferase | 0.85 | 1.22 | 1.12 |
| x30500 | Microalbumin in urine | 0.66 | 0.47 | 1.02 |
| x1835 | Mother still alive | 0.57 | 0.67 | 0.97 |
| x2000 | Worry too long after embarrassment | 0.70 | 0.39 | 0.97 |
| x3062 | Forced vital capacity (FVC) | 0.44 | 0.50 | 0.95 |
| x30770 | IGF-1 | 0.80 | 0.28 | 0.93 |
| x400 | Time to complete round | 0.73 | 0.85 | 0.88 |
| x6138__2 | Qualifications - A levels/AS levels or equivalent | 0.68 | 0.17 | 0.78 |
| x131456 | Pneumonia, organism unspecified (J18) date | 0.69 | 0.57 | 0.77 |
| x23105 | Basal metabolic rate | 0.31 | 0.00 | 0.75 |
| x102 | Pulse rate, automated reading | 0.58 | 0.71 | 0.72 |
| x6138__3 | Qualifications - O levels/GCSEs or equivalent | 1.06 | 0.74 | 0.72 |
| x23107 | Impedance of leg (right) | 0.60 | 0.48 | 0.71 |
| x23111 | Leg fat percentage (right) | 0.09 | 0.17 | 0.69 |
| x41204__E780 | Pure hypercholesterolemia (E78.0) | 0.19 | 0.00 | 0.67 |
| x6159__100 | Pain type(s) experienced in last month - none | 0.58 | 0.45 | 0.64 |
| x3064 | Peak expiratory flow (PEF) | 0.33 | 0.45 | 0.61 |
| x30700 | Creatinine | 0.49 | 0.43 | 0.61 |
| x2316 | Wheeze or whistling in the chest in last year | 0.79 | 0.53 | 0.58 |
| x54 | UK Biobank assessment center | 0.64 | 0.46 | 0.57 |
| x21001 | Body mass index (BMI) | 0.31 | 0.37 | 0.55 |
| x137 | Number of treatments/medications taken | 0.43 | 0.55 | 0.53 |
| x41214__2 | Carer support indicators - no | 0.46 | 0.39 | 0.51 |
| x2267 | Use of sun/UV protection | 0.55 | 0.50 | 0.50 |
| x30690 | Cholesterol | 0.14 | 0.28 | 0.50 |
| x20023 | Mean time to correctly identify matches | 0.19 | 0.27 | 0.45 |
| x30750 | Glycated hemoglobin (HbA1c) | 0.49 | 0.41 | 0.44 |
| x1249 | Past tobacco smoking | 0.46 | 0.24 | 0.43 |
| x23120 | Arm fat mass (right) | 0.21 | 0.25 | 0.41 |
| x30670 | Urea | 0.25 | 0.22 | 0.38 |
| x41272__Z943 | Left sided operation (Z94.3) | 0.13 | 0.00 | 0.37 |
| x30650 | Aspartate aminotransferase | 0.17 | 0.27 | 0.35 |
| x399 | Number of incorrect matches in round | 0.46 | 0.36 | 0.34 |
| x680 | Own or rent accommodation lived in | 0.37 | 0.25 | 0.33 |
| x1883 | Number of full sisters | 0.35 | 0.21 | 0.32 |
| x30620 | Alanine aminotransferase | 0.21 | 0.20 | 0.32 |
| x4080 | Systolic blood pressure, automated reading | 0.24 | 0.22 | 0.31 |
| x20015 | Sitting height | 0.10 | 0.24 | 0.30 |
| x30510 | Creatinine (enzymatic) in urine | 0.29 | 0.27 | 0.30 |
| x30520 | Potassium in urine | 0.13 | 0.11 | 0.30 |
| x1488 | Tea intake | 0.25 | 0.26 | 0.29 |
| x30880 | Urate | 0.17 | 0.35 | 0.29 |
| x21000 | Ethnic background | 0.20 | 0.39 | 0.28 |
| x30610 | Alkaline phosphatase | 0.29 | 0.20 | 0.28 |
| x30840 | Total bilirubin | 0.17 | 0.20 | 0.28 |
| x1408 | Cheese intake | 0.30 | 0.05 | 0.27 |
| x50 | Standing height | 0.20 | 0.14 | 0.27 |
| x6164__5 | Heavy DIY physical activity in last 4 weeks | 0.25 | 0.12 | 0.27 |
| x132070 | Other disorders of urinary system (N39) date | 0.24 | 0.07 | 0.26 |
| x943 | Frequency of stair climbing in last 4 weeks | 0.17 | 0.43 | 0.26 |
| x41204__Z864 | Personal history of psychoactive substance abuse (Z86.4) | 0.08 | 0.08 | 0.24 |
| x699 | Length of time at current address | 0.19 | 0.44 | 0.24 |
| x52 | Month of birth | 0.16 | 0.08 | 0.23 |
| x6154__100 | Medication for pain relief, constipation, heartburn - none | 0.17 | 0.20 | 0.23 |
| x884 | Number of days/week of moderate physical activity 10+ minutes | 0.21 | 0.13 | 0.23 |
| x1438 | Bread intake | 0.12 | 0.25 | 0.22 |
| x20002__1065 | Non-cancer illness code, self-reported - hypertension | 0.27 | 0.00 | 0.21 |
| x130814 | Disorders of lipoprotein metabolism and other lipedemas (E78) date | 0.17 | 0.31 | 0.20 |
| x135 | Number of self-reported non-cancer illnesses | 0.08 | 0.00 | 0.19 |
| x1538 | Major dietary changes in the last 5 years | 0.06 | 0.00 | 0.19 |
| x41204__E119 | Type 2 diabetes mellitus without complications (E11.9) | 0.16 | 0.13 | 0.19 |
| x1727 | Ease of skin tanning | 0.24 | 0.14 | 0.17 |
| x41270__J181 | Lobar pneumonia, unspecified (J18.1) | 0.21 | 0.17 | 0.17 |
| x47 | Hand grip strength (right) | 0.08 | 0.20 | 0.17 |
| x41272__G451 | Fiberoptic endoscopic examination (G45.1) | 0.08 | 0.08 | 0.16 |
| x1498 | Coffee intake | 0.13 | 0.14 | 0.15 |
| x30530 | Sodium in urine | 0.15 | 0.14 | 0.15 |
| x41204__F329 | Depressive episode, unspecified (F32.9) | 0.12 | 0.05 | 0.15 |
| x136 | Number of operations, self-reported | 0.08 | 0.06 | 0.13 |
| x1418 | Milk type used | 0.12 | 0.19 | 0.13 |
| x20116 | Smoking status | 0.06 | 0.09 | 0.13 |
| x2040 | Risk taking | 0.19 | 0.13 | 0.12 |
| x4079 | Diastolic blood pressure, automated reading | 0.18 | 0.23 | 0.12 |
| x1647 | Country of birth (UK/elsewhere) | 0.14 | 0.12 | 0.11 |
| x1747 | Hair color (natural, before greying) | 0.14 | 0.11 | 0.11 |
| x6164__100 | Types of physical activity in last 4 weeks - none | 0.12 | 0.09 | 0.11 |
| x131492 | Other chronic obstructive pulmonary disease (J44) date | 0.07 | 0.11 | 0.10 |
| x6138__1 | Qualifications - college or university degree | 0.17 | 0.15 | 0.10 |
| x6146__100 | Attendance/disability/mobility allowance - none | 0.07 | 0.08 | 0.10 |
| x1458 | Cereal intake | 0.08 | 0.07 | 0.09 |
| x41270__D649 | Anemia, unspecified (D64.9) | 0.10 | 0.08 | 0.09 |
| x49 | Hip circumference | 0.16 | 0.06 | 0.09 |
| x1070 | Time spent watching television (TV) | 0.30 | 0.11 | 0.08 |
| x130648 | Other anemias (D64) date | 0.08 | 0.02 | 0.08 |
| x1448 | Bread type | 0.08 | 0.14 | 0.08 |
| x1478 | Salt added to food | 0.12 | 0.10 | 0.08 |
| x41272__U051 | Computed tomography of head (U5.1) | 0.10 | 0.03 | 0.08 |
| x904 | Number of days/week of vigorous physical activity 10+ minutes | 0.15 | 0.05 | 0.08 |
| x1797 | Father still alive | 0.22 | 0.05 | 0.07 |
| x21002 | Weight | 0.10 | 0.15 | 0.07 |
| x41204__Z867 | Personal history of diseases of the circulatory system (Z86.7) | 0.06 | 0.07 | 0.07 |
| x41270__R11 | Nausea and vomiting (R11) | 0.08 | 0.05 | 0.07 |
| x6155__100 | Vitamin and mineral supplements - none | 0.07 | 0.00 | 0.07 |
| x1319 | Dried fruit intake | 0.26 | 0.04 | 0.06 |
| x1389 | Pork intake | 0.06 | 0.07 | 0.06 |
| x20003__1141188442 | Glucosamine product | 0.06 | 0.00 | 0.06 |
| x20004__1439 | Operation - reduction or fixation of bone fracture | 0.05 | 0.00 | 0.06 |
| x30870 | Triglycerides | 0.14 | 0.06 | 0.06 |
| x6145__6 | Illness, injury, bereavement, stress in last 2 years - financial difficulties | 0.06 | 0.15 | 0.06 |
| x1349 | Processed meat intake | 0.07 | 0.00 | 0.05 |
| x1369 | Beef intake | 0.06 | 0.00 | 0.05 |
| x2473 | Other serious medical condition/disability diagnosed by doctor | 0.11 | 0.05 | 0.05 |
| x40008 | Age at cancer diagnosis | 0.06 | 0.03 | 0.04 |
| x6145__100 | Illness, injury, bereavement, stress in last 2 years - none | 0.06 | 0.09 | 0.04 |
| x670 | Type of accommodation lived in | 0.06 | 0.11 | 0.04 |
| x1873 | Number of full brothers | 0.08 | 0.03 | 0.03 |
| x1940 | Irritability | 0.08 | 0.03 | 0.02 |
| x132146 | Excessive, frequent and irregular menstruation (N92) date | 0.05 | 0.00 | 0.00 |
| x1980 | Worrier / anxious feelings | 0.11 | 0.00 | 0.00 |
| x20004__1458 | Operation - appendicectomy | 0.10 | 0.07 | 0.00 |
| x20110__5 | Illnesses of mother - breast cancer | 0.07 | 0.00 | 0.00 |
| x2335 | Chest pain or discomfort | 0.07 | 0.00 | 0.00 |
| x6149__4 | Mouth/teeth dental problems - loose teeth | 0.08 | 0.03 | 0.00 |
| x6152__100 | Blood clot, DVT, bronchitis, emphysema, asthma, rhinitis, eczema, allergy - none | 0.09 | 0.08 | 0.00 |
| x6154__3 | Medication for pain relief, constipation, heartburn - paracetamol | 0.05 | 0.08 | 0.00 |
