## supplementary table 5 for "Identifying risk factors for COVID-19 severity and mortality in the UK Biobank"

**Supplementary Table 5**. Listing of the important features identified using SHAP values for COVID-19 mortality models. Three sets of SHAP values are provided: a) SHAP values from GBDT models with all the features in their input totalling 62%, b) SHAP values from GBDT models using only the important features (SHAP value > 0.1% when all the features were input), and c) SHAP values from GBDT models with only the important features in their input, after removing 40 highly correlated features (correlation above 0.9)

| **Feature ID** | **Description** | **SHAP value (all features)** | **SHAP value (important features)** | **SHAP value (after removing highly correlated features)** |
| --- | --- | --- | --- | --- |
| x21022 | Age at recruitment | 6.56 | 4.56 | 13.52 |
| x4080 | Systolic blood pressure, automated reading | 1.77 | 2.12 | 2.59 |
| x30770 | IGF-1 | 1.04 | 1.47 | 1.58 |
| x48 | Waist circumference | 1.23 | 1.12 | 1.54 |
| x400 | Time to complete round | 0.68 | 1.23 | 1.46 |
| x1329 | Oily fish intake | 0.89 | 0.98 | 1.38 |
| x47 | Hand grip strength (right) | 1.05 | 0.95 | 1.38 |
| x30670 | Urea | 0.79 | 1.15 | 1.28 |
| x102 | Pulse rate, automated reading | 0.80 | 1.08 | 1.25 |
| x189 | Townsend deprivation index at recruitment | 0.93 | 1.06 | 1.21 |
| x30610 | Alkaline phosphatase | 0.78 | 1.07 | 1.21 |
| x399 | Number of incorrect matches in round | 0.83 | 1.05 | 1.18 |
| x23109 | Impedance of arm (right) | 0.55 | 0.90 | 1.17 |
| x20002__1065 | Non-cancer illness code, self-reported - hypertension | 0.61 | 0.29 | 1.13 |
| x30650 | Aspartate aminotransferase | 0.52 | 0.71 | 1.09 |
| x30840 | Total bilirubin | 0.89 | 1.03 | 1.04 |
| x1873 | Number of full brothers | 0.78 | 1.04 | 1.04 |
| x4079 | Diastolic blood pressure, automated reading | 0.59 | 0.61 | 1.02 |
| x1070 | Time spent watching television (TV) | 0.55 | 0.61 | 1.01 |
| x30720 | Cystatin C | 0.39 | 0.77 | 1.00 |
| x884 | Number of days/week of moderate physical activity 10+ minutes | 0.34 | 0.43 | 0.58 |
| x31 | Sex | 0.12 | 0.05 | 0.56 |
| x30700 | Creatinine | 0.39 | 0.39 | 0.56 |
| x41270__R69 | Unknown and unspecified causes of morbidity (R69) | 0.30 | 0.38 | 0.56 |
| x52 | Month of birth | 0.28 | 0.51 | 0.56 |
| x6149__1 | Mouth/teeth dental problems - mouth ulcers | 0.36 | 0.46 | 0.55 |
| x41200__G459 | Fiberoptic endoscopic examination of upper gastrointestinal tract (G45.9) | 0.24 | 0.38 | 0.53 |
| x23120 | Arm fat mass (right) | 0.30 | 0.38 | 0.52 |
| x2010 | Suffer from 'nerves' | 0.56 | 0.49 | 0.51 |
| x41204__E119 | Type 2 diabetes mellitus without complications (E11.9) | 0.14 | 0.31 | 0.47 |
| x41204__F329 | Depressive episode, unspecified (F32.9) | 0.27 | 0.53 | 0.46 |
| x1349 | Processed meat intake | 0.33 | 0.60 | 0.46 |
| x21002 | Weight | 0.48 | 0.39 | 0.45 |
| x2227 | Other eye problems | 0.23 | 0.36 | 0.45 |
| x23105 | Basal metabolic rate | 0.17 | 0.13 | 0.44 |
| x1428 | Spread type | 0.21 | 0.38 | 0.44 |
| x30500 | Microalbumin in urine | 0.17 | 0.35 | 0.42 |
| x2345 | Ever had bowel cancer screening | 0.29 | 0.28 | 0.41 |
| x41272__G451 | Fiberoptic endoscopic examination (G45.1) | 0.12 | 0.31 | 0.41 |
| x41270__M179 | Gonarthrosis, unspecified (M17.9) | 0.24 | 0.36 | 0.40 |
| x2306 | Weight change compared with 1 year ago | 0.14 | 0.16 | 0.39 |
| x1687 | Comparative body size at age 10 | 0.20 | 0.21 | 0.38 |
| x1379 | Lamb/mutton intake | 0.19 | 0.30 | 0.37 |
| x40008 | Age at cancer diagnosis | 0.22 | 0.37 | 0.37 |
| x131416 | Hypotension (I95) date | 0.20 | 0.36 | 0.37 |
| x728 | Number of vehicles in household | 0.23 | 0.40 | 0.36 |
| x2188 | Long-standing illness, disability or infirmity | 0.10 | 0.18 | 0.36 |
| x1289 | Cooked vegetable intake | 0.11 | 0.13 | 0.35 |
| x20003__1140864752 | Treatment/medication - lansoprazole | 0.16 | 0.23 | 0.35 |
| x924 | Usual walking pace | 0.17 | 0.36 | 0.34 |
| x6149__6 | Mouth/teeth dental problems - dentures | 0.20 | 0.31 | 0.34 |
| x1319 | Dried fruit intake | 0.26 | 0.35 | 0.34 |
| x1930 | Miserableness | 0.22 | 0.29 | 0.33 |
| x131876 | Other arthrosis (M19) date | 0.14 | 0.16 | 0.33 |
| x680 | Own or rent accommodation lived in | 0.12 | 0.25 | 0.32 |
| x6139__1 | A gas hob or gas cooker for cooking/heating | 0.17 | 0.27 | 0.32 |
| x41200__H229 | Unspecified diagnostic endoscopic examination of colon(H22.9) | 0.28 | 0.32 | 0.31 |
| x1299 | Salad / raw vegetable intake | 0.13 | 0.49 | 0.31 |
| x6164__2 | Physical activity in last 4 weeks - other exercises | 0.35 | 0.30 | 0.31 |
| x41272__Z942 | Right sided operation (Z94.2) | 0.12 | 0.27 | 0.31 |
| x135 | Number of self-reported non-cancer illnesses | 0.21 | 0.27 | 0.30 |
| x41270__K573 | Diverticular disease of large intestine without perforation or abscess (K57.3) | 0.12 | 0.24 | 0.29 |
| x1528 | Water intake | 0.16 | 0.22 | 0.28 |
| x136 | Number of operations, self-reported | 0.19 | 0.19 | 0.28 |
| x1389 | Pork intake | 0.12 | 0.31 | 0.27 |
| x41202__R074 | Chest pain, unspecified (R7.4) | 0.11 | 0.32 | 0.27 |
| x6154__2 | Medication for pain relief, constipation, heartburn - ibuprofen | 0.31 | 0.40 | 0.26 |
| x2040 | Risk taking | 0.11 | 0.19 | 0.25 |
| x131386 | Other peripheral vascular diseases (I73) date | 0.12 | 0.27 | 0.25 |
| x20003__99999 | Treatment/medication code - free-text, unable to be coded | 0.17 | 0.22 | 0.25 |
| x6139__3 | An open solid fuel fire that you use regularly in winter time for cooking/heating | 0.28 | 0.19 | 0.25 |
| x6179__1 | Mineral and other dietary supplements - fish oil | 0.14 | 0.21 | 0.24 |
| x1883 | Number of full sisters | 0.14 | 0.11 | 0.23 |
| x20002__1473 | Non-cancer illness code, self-reported - high cholesterol | 0.17 | 0.16 | 0.23 |
| x6155__7 | Vitamin and mineral supplements - multivitamins +/- minerals | 0.14 | 0.15 | 0.22 |
| x131872 | gonarthrosis [arthrosis of knee] (M17) date | 0.20 | 0.20 | 0.22 |
| x2247 | Hearing difficulty/problems | 0.15 | 0.18 | 0.22 |
| x6152__8 | Asthma diagnosed by doctor | 0.18 | 0.17 | 0.22 |
| x1970 | Nervous feelings | 0.22 | 0.17 | 0.22 |
| x20003__1140879802 | Treatment/medication - amlodipine | 0.24 | 0.19 | 0.22 |
| x1418 | Milk type used | 0.25 | 0.30 | 0.21 |
| x1950 | Sensitivity / hurt feelings | 0.14 | 0.16 | 0.21 |
| x131464 | vasomotor and allergic rhinitis (J30) date | 0.16 | 0.26 | 0.21 |
| x20002__1387 | Non-cancer illness code, self-reported - hayfever/allergic rhinitis | 0.14 | 0.10 | 0.21 |
| x6145__2 | Serious illness, injury or assault of a close relative in last 2 years | 0.22 | 0.26 | 0.21 |
| x6159__100 | Pain type(s) experienced in last month - none | 0.15 | 0.07 | 0.19 |
| x1990 | Tense / 'highly strung' | 0.16 | 0.15 | 0.19 |
| x41210__Y973 | Radiology with post contrast (Y97.3) | 0.13 | 0.30 | 0.19 |
| x6159__4 | Pain type(s) experienced in last month - back pain | 0.12 | 0.13 | 0.17 |
| x1239 | Current tobacco smoking | 0.11 | 0.11 | 0.17 |
| x20004__1510 | Operation - dilatation and curettage/d+c | 0.13 | 0.14 | 0.16 |
| x6162__2 | Types of transport used (excluding work) - walk | 0.16 | 0.11 | 0.16 |
| x3088 | Contra-indications for spirometry | 0.12 | 0.10 | 0.16 |
| x2257 | Hearing difficulty/problems with background noise | 0.13 | 0.14 | 0.16 |
| x670 | Type of accommodation lived in | 0.18 | 0.07 | 0.15 |
| x41270__N179 | Acute renal failure, unspecified (N17.9) | 0.12 | 0.14 | 0.14 |
| x41202__N390 | Urinary tract infection, site not specified (N39) | 0.13 | 0.14 | 0.14 |
| x1717 | Skin color | 0.13 | 0.11 | 0.13 |
| x131954 | Shoulder lesions (M75) date | 0.13 | 0.08 | 0.13 |
| x2277 | Frequency of solarium/sunlamp use | 0.14 | 0.13 | 0.12 |
| x6154__1 | Medication for pain relief, constipation, heartburn - aspirin | 0.11 | 0.04 | 0.11 |
| x2060 | Frequency of unenthusiasm / disinterest in last 2 weeks | 0.18 | 0.08 | 0.11 |
| x20004__1403 | Operation - inguinal/femoral hernia repair | 0.11 | 0.08 | 0.11 |
| x41200__U051 | Computed tomography of head (U5.1) | 0.12 | 0.11 | 0.11 |
| x41210__Y981 | Radiology of one body area (or < 2 minutes) (Y98.1) | 0.15 | 0.12 | 0.10 |
| x1707 | Handedness (chirality/laterality) | 0.13 | 0.13 | 0.10 |
| x131402 | Varicose veins of lower extremities (I83) date | 0.12 | 0.11 | 0.08 |
| x6164__4 | Light DIY physical activity in last 4 weeks | 0.10 | 0.08 | 0.03 |
| x40009 | Reported occurrences of cancer | 0.11 | 0.05 | 0.00 |
| x699 | Length of time at current address | 1.20 | 1.54 | 1.88 |
| x41200__X998 | No procedure performed (X99.8) | 0.83 | 0.87 | 1.85 |
| x1835 | Mother still alive | 1.17 | 1.58 | 1.82 |
| x20023 | Mean time to correctly identify matches | 0.83 | 1.15 | 1.78 |
| x30730 | Gamma glutamyl transferase | 1.38 | 1.83 | 1.64 |
| x6159__6 | Pain type(s) experienced in last month - hip pain | 0.58 | 0.71 | 0.60 |
| x21001 | Body mass index (BMI) | 0.44 | 0.47 | 0.90 |
| x41270__N390 | Urinary tract infection, site not specified (N39) | 0.33 | 0.84 | 0.91 |
| x30510 | Creatinine (enzymatic) in urine | 0.57 | 0.62 | 0.62 |
| x120 | Birth weight not known | 0.62 | 0.85 | 0.87 |
| x943 | Frequency of stair climbing in last 4 weeks | 0.31 | 0.49 | 0.62 |
| x1488 | Tea intake | 0.72 | 0.86 | 0.94 |
| x30750 | Glycated hemoglobin (HbA1c) | 0.75 | 0.76 | 0.94 |
| x23107 | Impedance of leg (right) | 0.47 | 0.70 | 0.96 |
| x6150__100 | Vascular/heart problems diagnosed by doctor - none | 0.37 | 0.64 | 0.64 |
| x30520 | Potassium in urine | 0.57 | 0.80 | 0.95 |
| x2267 | Use of sun/UV protection | 0.51 | 0.63 | 0.64 |
| x30620 | Alanine aminotransferase | 0.60 | 0.62 | 0.90 |
| x2000 | Worry too long after embarrassment | 0.41 | 0.60 | 0.72 |
| x30710 | C-reactive protein | 0.54 | 0.82 | 0.78 |
| x30870 | Triglycerides | 0.71 | 0.71 | 0.79 |
| x30530 | Sodium in urine | 0.66 | 0.74 | 0.93 |
| x23099 | Body fat percentage | 0.21 | 0.43 | 0.70 |
| x54 | UK Biobank assessment center | 0.34 | 0.44 | 0.69 |
| x49 | Hip circumference | 0.23 | 0.53 | 0.62 |
| x30880 | Urate | 0.77 | 0.91 | 0.84 |
| x41204__I10 | Essential (primary) hypertension (I10) | 0.56 | 0.44 | 0.60 |
| x30690 | Cholesterol | 0.65 | 0.52 | 0.92 |
| x20015 | Sitting height | 0.39 | 0.73 | 0.77 |
| x23128 | Trunk fat mass | 0.46 | 0.39 | 0.70 |
| x1438 | Bread intake | 0.47 | 0.60 | 0.63 |
| x1249 | Past tobacco smoking | 0.71 | 0.84 | 0.87 |
| x1448 | Bread type | 0.36 | 0.57 | 0.68 |
| x1339 | Non-oily fish intake | 0.30 | 0.36 | 0.67 |
| x50 | Standing height | 0.34 | 0.60 | 0.69 |
| x864 | Number of days/week walked 10+ minutes | 0.65 | 0.88 | 0.94 |
| x2080 | Frequency of tiredness / lethargy in last 2 weeks | 0.34 | 0.58 | 0.75 |
| x2492 | Taking other prescription medications | 0.71 | 0.66 | 0.85 |
| x6142__2 | Current employment status - retired | 0.34 | 0.35 | 0.64 |
| x1458 | Cereal intake | 0.99 | 0.96 | 0.96 |
